# Wellness in work - supporting people in work and assisting people to return to the workforce: An economic evidence review

**DOI:** 10.1101/2024.01.17.23300197

**Authors:** Rhiannon Tudor Edwards, Llinos Haf Spencer, Bethany Fern Anthony, Jacob Davies, Kalpa Pisavadia, Abraham Makanjuola, Huw Lloyd-Williams, Deborah Fitzsimmons, Brendan Collins, Joanna Mary Charles, Ruth Lewis, Alison Cooper, Sezen Barutcu, Mary-Ann McKibben, Adrian Edwards

## Abstract

Rapid review methodology was used to identify updated economic evidence on programmes or interventions designed to enable employees to remain in and return to the workforce. In Wales, there are currently 1.48 million people in employment and 58,300 people who are unemployed. This equates to an unemployment rate of 3.8% in Wales.

The evidence in this report builds on a previous [Wellness in Work](https://cheme.bangor.ac.uk/documents/Wellness-in-Work-Report.pdf) report (Edwards et al., 2019). This review has a broad focus to understand the economic impact of well-being within the workplace. The main section of the report is on the economic benefits of keeping the workforce well. Seventy-six papers were included from databases searched for literature published between 2017 and 2023.

Economics studies were found relating to common mental health conditions; severe mental health conditions; influenza vaccination; illicit drug use; smoking and vaping; healthy eating and physical activity. A range of economic evidence of low, medium, and high quality relating to interventions targeting well-being in the workforce was identified. This included high quality evidence that interventions in the workforce for employees at risk of common mental health disorders can be cost saving for businesses and the health and social care sector. There is also high quality evidence on the cost-effectiveness of interventions focusing on healthy eating and physical activity in the workplace. Some evidence gaps were also identified.

**Policy and practice implications:** There is a need to consider the evidence presented in this rapid review on cost-effective interventions to improve the wellness of the workforce in Wales. Updated policies and procedures to improve equal employment opportunities, regardless of age, gender, or disability status are needed.

**Economic considerations:** Amongst the G7 nations, the United Kingdom (UK) is performing relatively poorly in relation to returning to pre-pandemic employment rates. This is in part caused by the long elective (planned) surgery waiting lists present in the National Health Service (NHS) right across the UK, highlighting the circular relationship between health and the economy.

## 2. Foreword

The workforce landscape of Wales is changing rapidly due to demographic change, social and economic shocks such as the COVID-19 pandemic, Brexit and the cost-of-living crisis, changes to work patterns such as hybrid working and home working, increased use of meetings online, plus extra pressures on families due to stretched health and social care systems. Wales has a workforce of approximately 1.48 million, a fall from 1.5 million in 2019. Work is a central part of many of our lives defining who we are and to a great extent, determining how we spend our time, and integral to how we meet our caring responsibilities. Throughout the life-course we, as a society, spend the least amount on the health and social care of the population through these working years. These are the years that we are paying our taxes. It makes sense to prevent ill-health and promote well-being of the workforce. There are many benefits to being employed and these benefits include more spending power, being able to afford a good quality of life, being able to choose where to live geographically and contributing to wider society. Not all work is good work. Fair work environments, policies and practices can impact both positively (e.g., sense of purpose, social contact, supportive environment) and negatively (e.g., poor working conditions, unrealistic workloads, poor communications or relationships) on health, and work can cause or exacerbate health conditions (e.g., mental health and musculoskeletal issues - the two biggest causes of long-term sickness absence). There is a need for evidence on what are the most cost-effective ways of spending limited resources to enable people to stay well in work and manage the demands upon health and care services. Focusing on well-being in the workplace, this report provides evidence on relevant economic evaluations of interventions to support the health and well-being of the workforce across Wales.

**Professor Rhiannon Tudor Edwards**

Co-Director, Centre for Health Economics and Medicines Evaluation (CHEME), Bangor University

## 3. Executive summary

The evidence presented in this report builds on the 2019 [‘*Wellness in Work*’](https://cheme.bangor.ac.uk/documents/Wellness-in-Work-Report.pdf) report (Edwards et al., 2019). A rapid review methodology was used to identify updated economic evidence on programmes or interventions designed to enable employees to remain in and return to the workforce. In Wales, there are currently 1.48 million people in employment and 58,300 people who are unemployed. This equates to an unemployment rate of 3.8% in Wales.

This report has a broad focus to understand the economic impact of well-being within the workplace. The opening section of our report introduces the concept of well-being and discusses employment, unemployment and worklessness in Wales as well as self-employment and different types of employers. There are also sections on young people in work, old people in work, women in work, and a section about the health and social care workforce.

The main section in the report is about the economic benefits of keeping the workforce well. It focuses on the findings from the rapid searching of the worldwide literature of economic evidence for workplace programmes and interventions addressing common mental health difficulties, severe mental health difficulties, smoking cessation, controlling illicit drug use, healthy eating and physical activity in the workforce.

To be included in the rapid review, studies needed to report on economic evaluations, return on investment analyses, costing analyses, or work-related outcomes of economic interest (i.e., employment rates, sickness absence, presenteeism, work productivity). The searches were broad to capture a wide range of different workplace interventions. The search strategy included the terms ‘workplace’, ‘wellness at work’, ‘healthy workplace’ and health economics terms to capture economic evaluations and costing studies such as ‘cost-effectiveness’, ‘cost-benefit’ and ‘cost analyses’.

## 4. Lay Summary

Work is a central part of many of our lives defining who we are and to a great extent, determining how we spend our time, and vital to how we meet our caring responsibilities. Flexible working policies available to all staff can help people with caring responsibilities to re-join or maintain employment. The reduction of sickness-related absence could support people to remain in the workforce or return to the workforce.

Staff well-being is an important factor in workplace productivity. Promoting employee mental well-being will lead to financial gains.

Programmes targeting people living with short and long-term disabilities, as well as neurodiverse people to join and stay in the workforce, could increase inclusivity, and bring economic benefits to the Welsh economy. Supporting older people to stay in the workforce for longer can generate productivity gains through retaining experience and skills in the workplace.

There is growing but mixed evidence of the cost-effectiveness of workplace interventions particularly in the areas of flexible working, addressing employee mental health, and encouraging healthy behaviours.

## 5. About this report

This *Wellness in work* report builds upon existing evidence in the academic and grey literature. This report is embedded in the United Kingdom (UK) policy landscape focusing on Wales concerning work (Chwarae Teg, 2022; Hickman et al., 2023; Public Health Wales, 2023b). See Table 1 for relevant guideline documents for keeping people well in work.

**Table 1.**
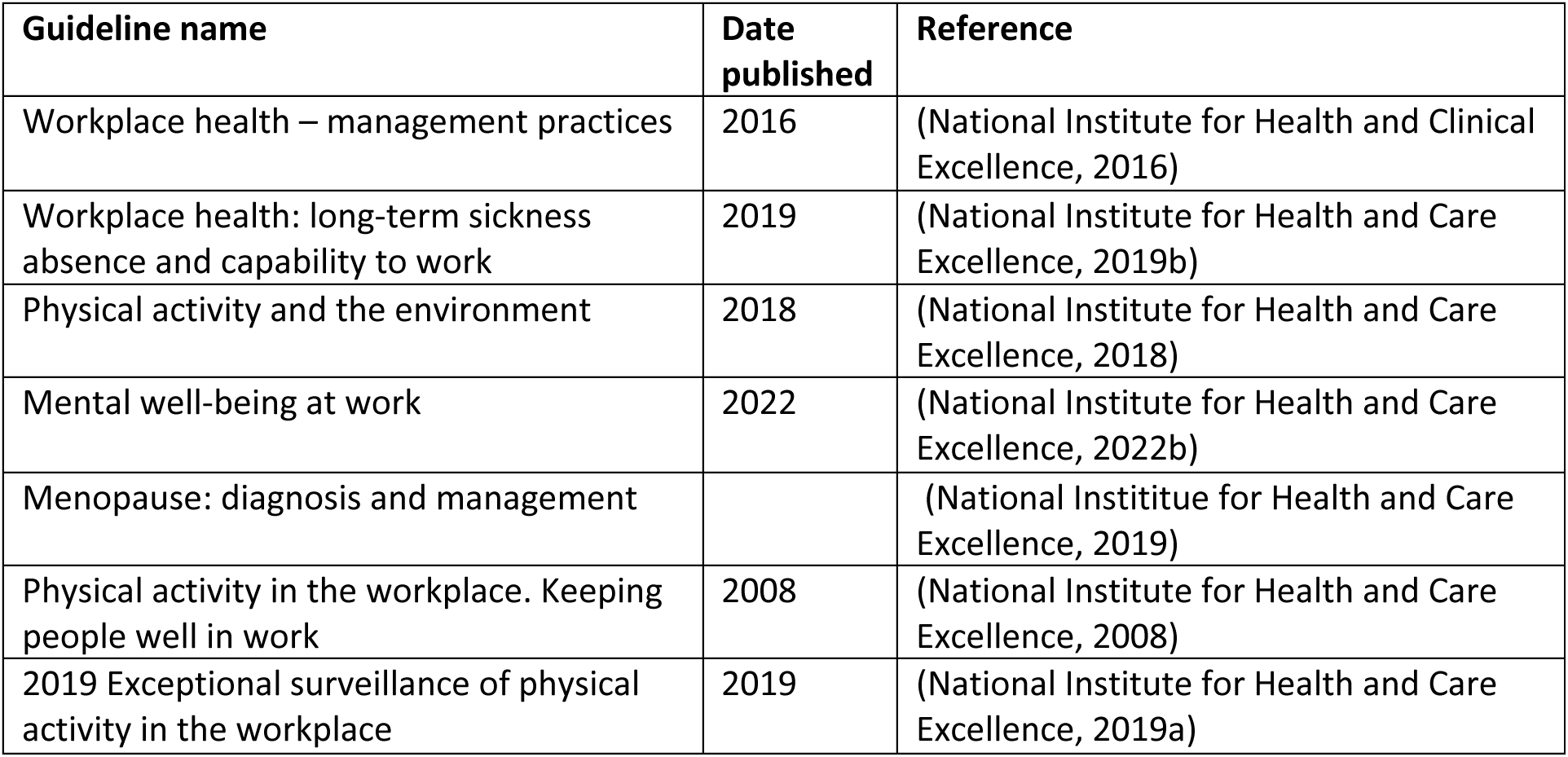
Relevant guideline documents for keeping people well in work.

| Guideline name | Date published | Reference |
| --- | --- | --- |
| Workplace health – management practices | 2016 | (National Institute for Health and Clinical Excellence, 2016) |
| Workplace health: long-term sickness absence and capability to work | 2019 | (National Institute for Health and Care Excellence, 2019b) |
| Physical activity and the environment | 2018 | (National Institute for Health and Care Excellence, 2018) |
| Mental well-being at work | 2022 | (National Institute for Health and Care Excellence, 2022b) |
| Menopause: diagnosis and management |  | (National Institute for Health and Care Excellence, 2019) |
| Physical activity in the workplace. Keeping people well in work | 2008 | (National Institute for Health and Care Excellence, 2008) |
| 2019 Exceptional surveillance of physical activity in the workplace | 2019 | (National Institute for Health and Care Excellence, 2019a) |

Some of the economic evidence presented in this report was not identified in our rapid review searches. We conducted supplementary literature searches for some of the main topics presented in this report to account for evidence on topics not picked up on in our searches. A glossary of terms is presented in Section 15 of this report.

## 6. Well-being

Whilst there is no consensus definition on what well-being is, it can be thought of as the balance point between an individual’s resource pool and the challenges faced (Dodge et al., 2012). Well-being needs to be nurtured to optimise health. Well-being is influenced by a myriad of factors across all life-course stages. Influences range from personal factors such as home and work environment; local factors such as access to health and social care, health literacy and justice to wider national and global factors such as social and cultural norms (Edwards et al., 2016). For this *Wellness in work* report, we use the *Well-being and well-becoming wheel* infographic (Edwards, 2022 - see Figure 1) to reflect how different personal, local, national, and global factors have an impact on well-being and well-becoming of individuals across through the life-course, including during people’s working lives.

**Figure 1:**
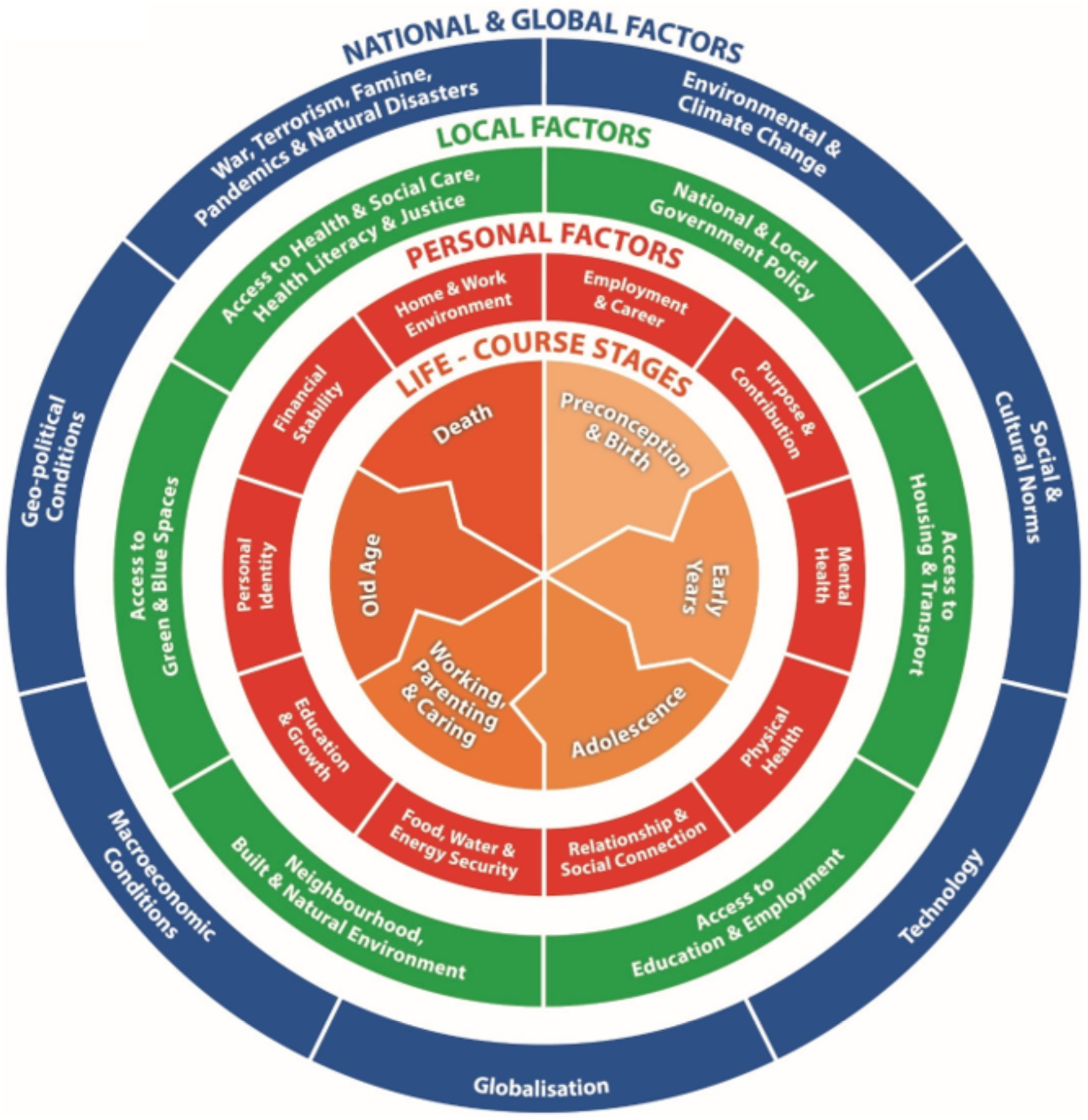
The well-being and well becoming wheel. (Edwards, 2022)

Improving workforce well-being became an increased focus of employers since the 1980s, with many Healthcare Trusts using health risk assessments, establishing on-site exercise classes and well-being sessions for their employees (National Health Service [NHS] England, 2023). In Wales, many employers aim to promote a good balance between work and home life and have promoted family friendly policies. For example, many workplaces now have a flexible working policy, and since the COVID-19 pandemic, many people follow a hybrid working pattern (Chwarae Teg, 2022). Increasing job satisfaction can improve mental health, increase well-being, and reduce absenteeism (Cao et al., 2022). Increased employee well-being is associated with lower levels of absenteeism and sick leave, greater resilience, better staff retention and increased staff commitment and productivity (MindWell, 2023).

### 6.1 Five ways to well-being

The five ways to well-being framework is used in parts of the National Health Service (NHS) in Wales (NHS, 2023). This framework suggests that there are five simple things that individuals can do on a daily basis to give their well-being a boost, such as: taking notice of their surroundings and savouring the moment; connecting with friends and family to build a sense of belonging and self-worth; being physically active to improve fitness and self-esteem; learning something new to increase self-confidence and sense of purpose, and giving to others, as acts of kindness, to create sense of reward (Figure 2).

**Figure 2:**
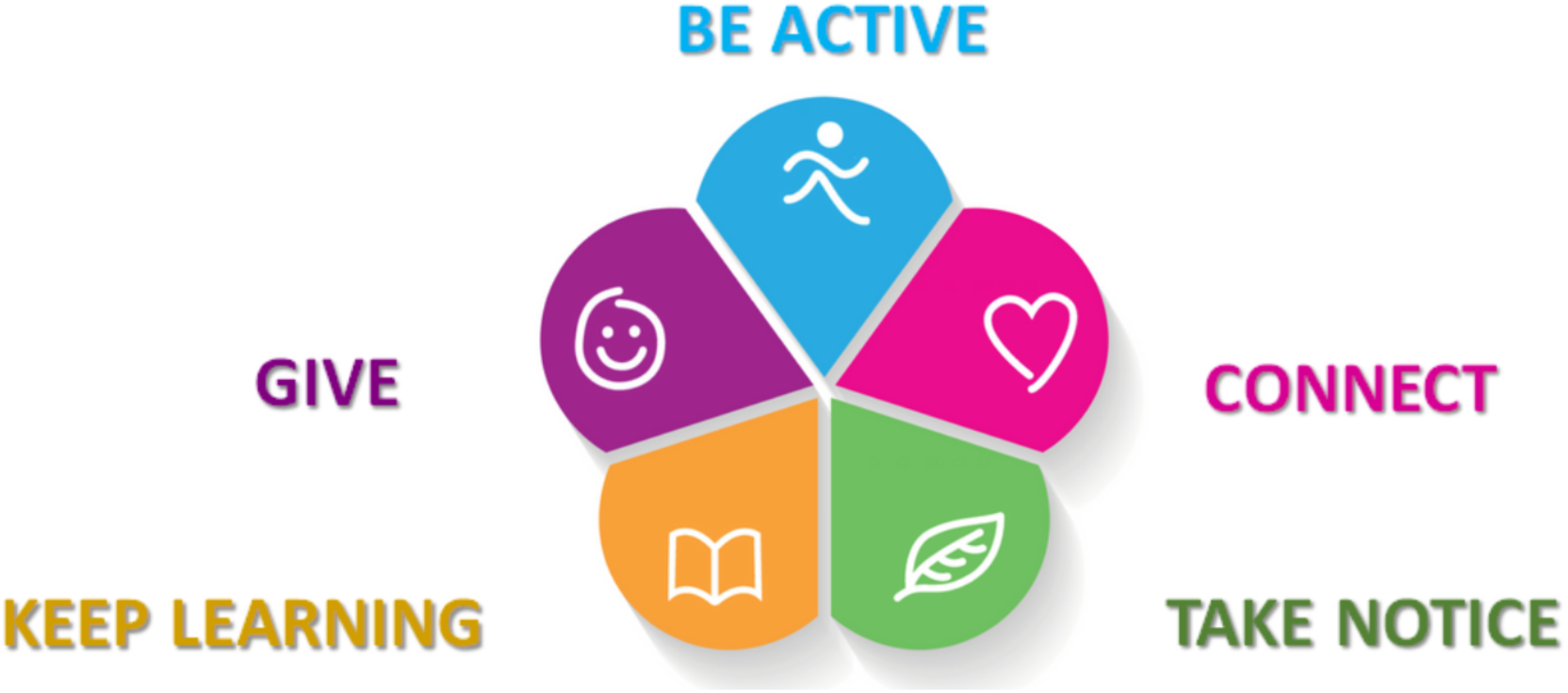
Five ways to well-being framework infographic. Source: Betsi Cadwaladr University Health Board. (Betsi Cadwaladr University Health Board, 2023)

## 7 Employment in Wales in the post-COVID-19 and post-Brexit era

There are currently 1.48 million working age individuals employed in Wales (74%). This is a decrease of 0.7 percentage points (PP) from the 75% employment rate in 2019 (Welsh Government, 2023g). here are 780,000 men in employment in Wales (79% of working-age men), an increase of 13,000 since 2019. There are 704,000 women in employment in Wales (69% of working-age women). Since 2019, 16,000 women have left employment in Wales. During the same period, 13,000 men have gained employment (Welsh Government, 2023g).

### 7.1 Economic activity and inactivity in Wales

Currently, 444,000 working age individuals are economically inactive in Wales: a rate of 23% (Welsh Government, 2023g). Economic inactivity has trended downward throughout 2023 but remains above the UK average. The Welsh economic inactivity rate has consistently been above the UK rate since 2019 (Welsh Government, 2023h). Of those economically inactive in Wales, 173,000 were men and 271,000 were women. The number of economically inactive men has fallen by 26,000 from 2022. The number of economically inactive women has also decreased in Wales since 2022, by 18,000. Indicating re-entry to the labour force for both sexes (Welsh Government, 2023g)(Welsh Government, 2023h).

The most common reasons for being economically inactive prior to reporting long-term sickness in the UK include looking after family or home (22%), being temporarily sick or injured (21%), retiring from paid work (18%) and being a student (12%) (Hickman et al., 2023).

At the UK level, increasing sickness patterns, changes in migration structures, retirement patterns and an ageing population contribute to rising economic inactivity (House of Lords Economic Affairs Committee, 2023). Dealing with preventable health issues, unhealthy behaviours, and reducing the risk of injuries may decrease premature mortality and keep people at work for longer (Hussein et al., 2016). The thriving at work framework suggests that all employees may need some support to thrive. Those who are struggling need targeted support, and those who are ill and possibly off work need tailored support to return to work (Farmer & Stevenson, 2017). Taking care of staff in environments that positively encourage greater well-being can, in the health and social care setting, help health and care staff to care better for other people.

### 7.2 Diversity and inclusivity in the workforce in Wales

The workforce is the people engaged in or available for work, either in a country or area or in a particular firm or industry. In Wales, there are currently 1.48 million people in employment and 52,300 people who are unemployed. The unemployment rate in Wales in 2023 is 3.8% (Welsh Government, 2023g). The number of paid employees has increased in recent years but fell during the COVID-19 pandemic. The number of paid employees returned to pre-pandemic levels in July 2021 and has remained above this level since then (Welsh Government, 2023g). The median gross weekly earnings for full-time adults working in Wales were £598 in April 2022 (93% of the UK average of £640) (Welsh Government, 2022a). Many families are just about managing (JAM) financially, and the current cost of living crisis is taking more people from Wales over the poverty line (Citizens Advice Bureau, 2023). In 2022, despite having one working adult in the household, 32% of children in Wales lived in poverty, an increase from 29% in 2019 (Welsh Government, 2019b, 2023d).

More individuals from ethnic minority communities are now in employment in Wales. The employment rate for working-age ethnic minorities in Wales was 68% in 2022, a 3PP increase from 2021. Comparatively, the employment rate of working-age white individuals was 74% in 2022, a 0.1PP increase on the previous year (Welsh Government, 2023h). The unemployment rate of working-age ethnic minorities was 7% in 2022 (a 3.9PP decrease from 2022). The unemployment rate for working-age white individuals was 3% in 2022 (a 1.1PP decrease from the previous year) (Welsh Government, 2023h).

Geographical location is strongly related to employment, and entrenched deprivation persists in Wales. High employment deprivation persisted in numerous areas between the 2014 and 2019 Welsh Index of Multiple Deprivation (WIMD), see Figure 3. The darker shaded regions in Figure 3 show areas categorised by level of employment deprivation (lack of employment). The key indicator for lack of employment used in the creation of the WIMD employment domain is the percentage of people in each area in receipt of employment-related benefits. High levels of employment deprivation exist in the South Wales valleys, large Welsh cities, and coastal towns in North Wales (Welsh Government, 2023o). The percentage of people in income deprivation in Lower Layer Super Output Areas (LSOAs) in deep-rooted deprivation (43%) was almost three times that of areas that have never been ranked in the top 50 most deprived (15%). It was also around three times the Wales average (16%) (Welsh Government, 2023o). Some communities are highly reliant on anchor institutions for their employment. Mass unemployment events can adversely impact the health, financial and social circumstances of workers, families, and communities (Davies et al., 2019). The recent closure of the 2 Sisters Food Group factory in Llangefni, Ynys Mon, saw the loss of 700 jobs in the local community (BBC Wales News, 2023).

**Figure 3:**
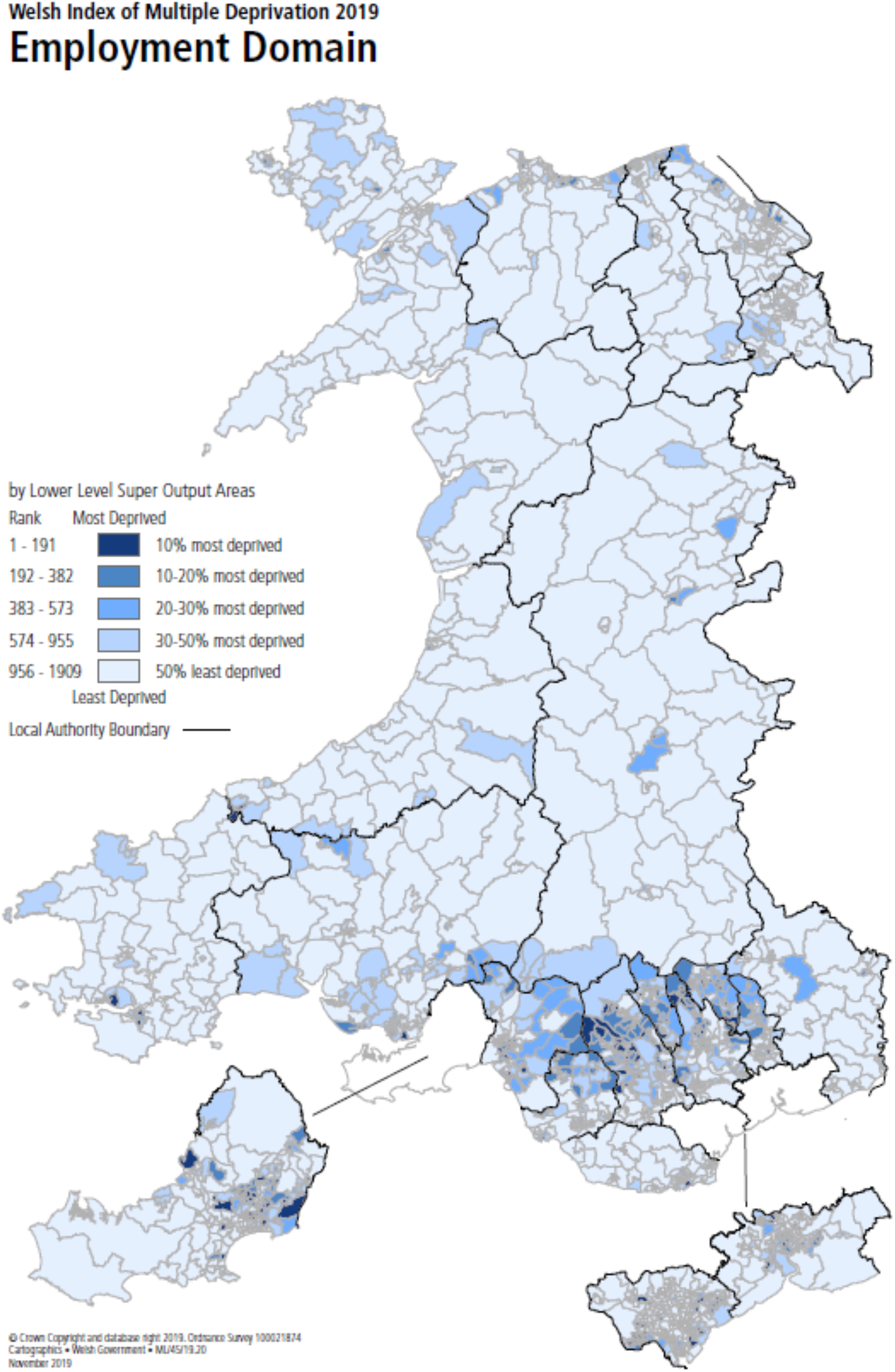
The areas most deprived in terms of employment in Wales. Source: Welsh Index of Multiple Deprivation (WIMD) 2019. (Welsh Government, 2023o)

### 7.3 The public sector continues to be a key employer within Wales

The public sector in Wales employed 324,000 people as of June 2023. This figure increased by 7,000 (2%) from 2022 (Welsh Government, 2023g). Human health and social work activities accounted for the greatest number of workforce jobs in Wales in 2022 at 223,000 (15%) of the total workforce (Office for National Statistics, 2023e). In 2019, the figure stood at 214,000, highlighting a longitudinal dependence on the sector in Wales.

### 7.4 Micro, small and medium sized enterprises

In 2022, micro, small and medium sized enterprises (SMEs) in Wales accounted for 63% of employment and 42% of turnover, with large enterprises accounting for the remainder. Most active enterprises were SMEs (10 to 249 employees), accounting for 99% of total enterprises in Wales. Micro enterprises (up to 9 employees) accounted for 95% of the total enterprises in Wales (Welsh Government, 2023m). The wholesale, retail, transport, hotels, food, and communication sectors had the largest proportion of enterprises and employment in SMEs, accounting for 25% of active SMEs in Wales and 31% of total employment (Welsh Government, 2023m). This has implications for supporting people with health problems, as occupational health support is often absent in SMEs.

#### 7.4.1 Gender

Overall, 13% of SME employers in Wales were women-led in 2019 (Department for Business Energy and Industrial Stragegy, 2020). The proportion is highest in small businesses (18%), lower at 13% in micro businesses and lower still (8%) in medium sized businesses (Department for Business Energy and Industrial Stragegy, 2020).

#### 7.4.2 Minority groups

Overall, 1.5% of SMEs in Wales were minority ethnic group-led businesses. The percentage is lowest for micro businesses (1.2%) and highest for medium sized businesses (3.4%) (Department for Business Energy and Industrial Stragegy, 2020).

### 7.5 Self-employment in Wales

In June 2023, 165,000 people were self-employed in Wales, which is 11.2% of the workforce (Welsh Government, 2023g). Many of these self-employed people are contractors, freelancers, and gig workers. Percentages of self-employed differ in each county of Wales. The average rate of self-employment across the counties of Wales is 12.4%, with percentages ranging from 5.6% in Blaenau Gwent and to 28% in Powys (Welsh Government, 2023f).

#### 7.5.1 Gender

In 2020, 8.8% of women and 16.9% of men in Wales were self-employed (Chwarae Teg, 2022).

#### 7.5.2 Minority groups

The rate of unemployment among ethnic minority women remains higher than among white women and is now higher than before the COVID-19 pandemic (Chwarae Teg, 2022).

### 7.6 The cost of living crisis in Wales

The UK has seen sharp rises in inflation since Winter 2021. Consumer Price Index including owner occupier housing costs (CPIH) rose from 3% in August 2021 to a peak of 9.6% in October 2022. CPIH is currently 6.3% (Office for National Statistics, 2023d). In the lead up to the cost of living crisis, Wales had the highest proportion of working-age adults (21%) and pensioners (18%) in relative income poverty out of the UK nations. 28% of children in Wales were living in relative poverty (Welsh Government, 2023o). Given that over half of all mental health problems start by age 14 (and 75% by age 18) and poverty being a known risk factor for psychological illnesses, there is likely to be a long shadow of mental health difficulties continuing into future generations (Department of Health, 2015; Fell & Hewstone, 2015).

The financial effects of the cost of living crisis is not felt equally. Poorer households tend to spend a greater proportion of their income on items more exposed to inflationary pressures, such as energy and food (Welsh Parliament, 2022). 90% of people in Wales felt their health was negatively affected due to increased heating costs (Royal College of Physicians, 2022). Decisions around the use of energy at home can lead to lingering anxiety that negatively impact mental health (Marmot, 2020).

## 8 Unemployment, worklessness and returning to work

### 8.1 Unemployment

In Wales, 58,000 individuals are currently unemployed: a rate of 3.8%. This has fallen from 62,000 in 2019 (Welsh Government, 2023f, 2023h). There are 36,000 unemployed men, an increase of 10,000 (40%) from 2022. This has remained almost static from the 36,000 level in 2019 (Welsh Government, 2023f). This increase may relate to a greater dependency on self-employment, a sector that has exhibited both volatility and a decline in number since 2020 (Office for National Statistics, 2023a). There are 17,000 women unemployed, a decrease of 3,000 (15%) from 2022. This decrease is a consistent trend observed from 2019, when 26,000 women were unemployed in Wales (Welsh Government, 2023f).

The Welsh Government has many strategies to help young people get into work and to support unemployed people to return work. These strategies include programmes such as Jobs Growth Wales+, a training and development programme for 16 to 19-year-olds that provides skills, qualifications, and experience that young people need to get a job or further training. Other programmes are not solely focused on young people, such as ReAct+. ReAct+ offers tailored solutions which may include financial support, skills training, and Personal Development Support to help remove barriers to employment, such as support with mental health, confidence building, language skills (Welsh Government, 2023k). Communities for Work Plus provides specialist employment advisory support and intensive mentoring to people who are under-represented in the labour market, including young, old, and disabled people; people from ethnic minority backgrounds (Welsh Government, 2023e).

### 8.2 Workless households

Workless households are households where no-one aged 16 or over is in employment. These members may be unemployed or economically inactive. Economically inactive members may be unavailable to work because of family commitments, retirement, study or unable to work through sickness or disability (Nomis, 2023).

> Well-being is essential for ensuring a healthy and productive labour force. Keeping people healthy and in work prevents loss of productivity and benefits the Welsh economy (Black, 2009).

## 9 Young people and paid employment

In 2023, it has been estimated that on the Annual Population Survey (APS) basis, the proportion of 16-18 year olds not in education, employment or training (NEET) was 10.5% in June 2023, compared with 6.3% for the year ending June 2022. When considering 19 to 24 year olds, 15.8% were NEET in June 2023 compared with 15.1% for the year ending June 2022 (Welsh Government, 2023q). NEETs often have diverse needs that require flexible and tailored solutions (Amendola, 2022; Nartey et al., 2014). Many young people do not have the necessary networking skills to assist in the job hunting process (Bonoli, 2014).

In recent years, there has been a growing focus on the mental health and well-being of young people in Wales (Neagle et al., 2018). For example, the ADTRAC programme was set up to assist young people into education, training, or work (Burgess et al., 2021). **A cost-benefit analysis on the ADTRAC programme showed a cost benefit ratio of £1.11 benefit for each £1 invested** (Burgess et al., 2021). A similar study called BackTrack conducted in a rural Australian setting found evidence of significant quantifiable improvements in several outcomes: high school attendance or completion, vocational education attendance or completion, unskilled or vocationally qualified employment and economic productivity as well as reduced homelessness, engagement with health services, acquisition of job readiness skills, as well as reduced local infrastructure vandalism and reduced crime (Deeming et al., 2022). In the BackTrack study, ‘high risk’ was defined as the increased likelihood of criminal activity, lack of employment, mental health issues, alcohol and substance abuse and lack of engagement with the health care system. Employment-related benefits of the intervention included educational attendance or completion of education, increased employment, and economic productivity. **The analysis of the BackTrack intervention produced a net social benefit of $3,267,967, with a return on investment (ROI) of $2.03 for every $1 invested** (Deeming et al., 2022). Both ADTRAC and BackTrack aim to address the needs of high-risk young people.

### 9.1 Apprenticeships

Apprenticeships are a way of introducing young people into the workforce. Apprenticeships in Wales are available to anyone aged 16 and over, including young people looking to transition from student life to working life, people who are unemployed, and people who have a job but are looking to switch careers. Although apprenticeships are not a novel intervention, they are encouraged by the Welsh Government through Career Wales with financial incentives for businesses to take on a learner in varying occupations, including construction, health, transport, retail, tourism, finance, electronics, and the media (Careers Wales, 2023; Welsh Government, 2023c). The number of learners who started apprenticeship learning programmes in 2022/23 was 7,170 compared with 7,560 starts in 2021/22 (Welsh Government, 2023b).

#### 9.1.1 Gender

The uptake of apprenticeships in Wales is equitable across genders. In 2022/23, around 48% of all apprenticeship learning programmes were taken up by women, compared with 42% in the previous year (Welsh Government, 2023b).

#### 9.1.2 Minority groups

In 2022/23, 355 (5%) learners who self-identified as being from ethnic minority backgrounds took up apprenticeships in Wales. This was an increase from 3.8% in 2021/22 (Welsh Government, 2023b).

## 10 Older people and paid employment

The default retirement age in the UK ended in 2011 to give people more choice about when to stop working (HM Revenue & Customs, 2021; Labour Market Reform, 2011). In March 2023, 657,600 people over the age of 65 were in employment. This is 8% of total employment in the UK, a decrease from a rate of 11% in 2019 (Office for National Statistics, 2023c). The state pension age changed to 66 in 2020 and is set to increase to 67 in March 2028. This gives many people no choice but to work past the age of 65 (HM Revenue & Customs, 2021).

Elective surgery waiting lists have reached 754,000 (or just under 1 in 4 of the total population of Wales). Age breakdowns are not available in the data, yet it is reasonable to argue a considerable amount of working age and older people are awaiting elective surgery that is limiting their ability to work or work productively (Welsh Government, 2023j).

Supporting older people to stay in the workforce for longer can generate productivity gains through retaining substantial amounts of experience and skills in the workplace (ILC, 2022). Women aged 50 to 74 living in the ‘healthiest’ areas of England and Wales were 6% more likely to be in paid work than those living in the ‘unhealthiest’ areas (ILC, 2022). Between the healthiest and unhealthiest areas of England and Wales, there is an 11-year gap in disability-free life expectancy (DFLE) (Office for National Statistics, 2020).

There is a gap in the evidence regarding productivity and older employees, but there is convincing evidence to suggest that worklessness may have detrimental effects on the well-being of an older person because of missing social connections, mental stimulation, confidence, being valued and making a positive contribution to society (Sewdas et al., 2017).

### 10.1.1 Gender

In the most deprived areas in Wales, women have an 18-year shorter healthy life expectancy than in the least deprived areas. This statistic for men is 17 years (The Health Foundation, 2022).

### 10.1.2 Minority groups

In terms of ethnicity, there was no difference in the employment rate between white people and people from ethnic minorities among 50- to 64-year-olds (71%) (there were no statistics for employees aged 65+ and ethnicity) (UK Population by ethnicity, 2020).

## 11 Women in paid employment

In December 2022, 9.7 million women were working full-time, and 5.9 million were working part-time in the UK. The total number of women working in the UK has increased by 1.7 million over the past 10 years (Buchanan et al., 2023). The employment rate for women is lower in Wales compared to the rest of the UK, and the gap between men and women has widened significantly from 0.8PP to 5.2PP in the year since 2022 (Welsh Government, 2022c).

### 11.1 The gender pay gap

The gender pay gap is currently 15% for all employees, primarily traced back to the fall in women’s average salary after becoming parents (Bari, 2023; Buchanan et al., 2023). Gender gaps in the labour market increase amongst employees in their late 20s and early 30s, suggesting that parenthood is potentially the cause (Andrew et al., 2012). While the average salary of men is unaffected by parenthood, the average salary of women sees little increase after parenthood (Bari, 2023).

### 11.2 Maternity pay

In the UK, the current basic statutory maternity and parental pay from April 2023 is £172.48 per week for 33 weeks (Benefits and Financial Support for Families, 2023). This equates to 47% of the National Living Wage (for a 35-hour week at the adult rate of £10.47 per hour) (The National Minimum Wage, 2023). The failure of maternity pay to keep up with the cost of living combined with the increase in household spending due to a new baby has the potential to cause financial hardship and stress (All Party Parliamentary Group, 2023).

The rate of mothers in employment has overtaken the employment rates of women and men without dependent children since 2017 (76%) (Office for National Statistics, 2021a). The reasons for an increase in the employment rates of mothers may be due to various aspects of support that have been introduced over the last 20 years. This includes shared parental leave in 2015, in which parents were provided with the legal right to share maternity leave entitlement (Benefits and Financial Support for Families, 2023). Additionally, the UK Government has implemented various childcare schemes across the UK (Get childcare: step by step, 2023). For example, in Wales, a total of 30 hours per week of early education and childcare is provided by Welsh Government for three and four-year-olds (Glover et al., 2018). These childcare schemes have been shown to support parents’ return to work and increase their working hours (Glover et al., 2018; Ruppanner et al., 2019).

### 11.3 Polycystic ovary syndrome

Polycystic ovary syndrome (PCOS) is the most common endocrine condition in women of reproductive age and affects approximately 6% to 10% of women of reproductive age (Wekker et al., 2020). It is often diagnosed in the women of childbearing age who are confronted with infertility (Costello et al., 2019). PCOS is often linked to a variety of health problems that affect physical and emotional well-being in the long-term (World Health Organisation (WHO), 2023).

In 2018, the Welsh Government launched the Period Dignity Strategic Action Plan which considered the link between periods and broader health issues, the environmental impact of disposable sanitary products, the impact on the workplace and on engagement in sport and culture (Welsh Government, 2021). Lack of knowledge about what counts as ‘normal’ in terms of menstrual health can lead to the late diagnosis of serious conditions such as PCOS, premenstrual dysphoric disorder (PMDD), gynaecological cancers, and endometriosis (Welsh Government, 2021).

Since 2018 the Welsh Government has invested approximately £9 million to ensure that children, young people, and those on low incomes have access to free period products, disseminated through schools, colleges, and communities in Wales.

### 11.4 Endometriosis

Another condition that affects girls and working age women is endometriosis. This is a condition that affects in 10 women where tissue like the lining of the womb grows elsewhere in the body, such as on the ovaries and in the fallopian tubes (All Party Parliamentary Group on Endemetriosis, 2020). Symptoms include lower abdomen and back pain, severe pain during periods, pain during sex, bowel and bladder symptoms, and fertility problems. The condition can significantly impact women’s lives, such as suboptimal educational and employment attainment and economic inactivity (All Party Parliamentary Group on Endemetriosis, 2020). Endometriosis costs the UK economy £8.2 billion a year in treatment, loss of work and health care costs (All Party Parliamentary Group on Endemetriosis, 2020).

In 2022 the Welsh Government Quality Statement for Women and Girls’ Health ensured that health boards would provide appropriate levels of diagnostic, therapeutic and surgical capacity to enable women who require interventions for health needs specific to women and girls, including menstrual and fertility care, endometriosis, and menopause, to receive care as close as possible to home without significant waits (Welsh Government, 2023j). See Box 1, for working women in Wales summary.

### 11.5 Menopause

Around 70% to 80% of women aged between 45 and 55 transition through menopause while still in work, and half of the menopausal women in the workplace find it challenging to cope with work during menopause (Floresco, 2023). Menopause diagnosis and management guidelines were developed by the National Institute for Health and Care Excellence (NICE)(National Instititue for Health and Care Excellence, 2019) and since then, both NHS Wales and Welsh Government have produced menopause policies (NHS Wales, 2021; Welsh Government, 2023a). Increases in the number of economically inactive women could be caused by, but not limited to, symptoms of menopause or because of higher levels of caring responsibilities than men (Hickman et al., 2023).

Some symptoms of menopause can last for several years and be physically and emotionally distressing, negatively impacting workplace relationships. Symptoms can include anxiety, reduced concentration, sleep disturbance, hot flushes, and heavy periods. These symptoms can be severe enough to result in extended periods of absence or leaving work entirely (Chartered Institute for Personel Development, 2019). The average salary of a 50 to 59-year-old woman in the UK in 2023 is £31,356 (gross pay) (Forbes Advisor, 2023).

Following advice and recommendations from the All-Wales Menopause Task and Finish Group in January 2023, the Welsh Government is committed to improving menopause care and support for women across Wales (Welsh Government, 2023a). Some of these recommendations include timely access to primary, secondary, and tertiary care that is culturally sensitive and inclusive, and access to a range of hormone replacement therapy (HRT) preparations (Welsh Government, 2023a). The [NICE menopause guideline includes recommendations on individualized care, diagnosis of perimenopause and menopause, information and advice, managing short-term menopausal symptoms, long-term benefits and risks of hormone replacement therapy, and diagnosing and managing premature ovarian insufficiency](https://www.nice.org.uk/guidance/NG23) (National Instititue for Health and Care Excellence, 2019)

##### Box 1 Working women in Wales summary

In 2023, 666,00 women are in employment in Wales (67% of working age women).

Both menopause and endometriosis have been identified as issues that either make women leave the workforce early or take many sickness-related days. Both conditions are challenging for women and their employers. Since 2015 there has been more emphasis by NICE, NHS Wales and the Welsh Government to support menopausal women in the workplace (National Institute for Health and Care Excellence, 2015; NHS Wales, 2021; Welsh Government, 2023a).

## 12 Disabled individuals in the workforce

People with disabilities are less represented in the workforce in the UK than non-disabled individuals. In 2022, 53% of people in the UK with a disability were in paid employment, significantly less than the 83% of non-disabled individuals (Department for Work and Pensions, 2023). For the year 2021/22, there were 462,000 disabled people in Wales. Of these, 227,000 were in paid employment, giving a disability employment rate of 49%, slightly lower than the UK average (Welsh Government, 2023h). The disability employment gap is a measure extensively used to compare the difference in employment rates of disabled individuals against individuals without disabilities (Powell et al., 2018). The disability employment gap is calculated from the difference between the non-disabled employment rate (81%) and the disability employment rate (49%), giving a disability employment gap for Wales in 2021/22 of 32.3PP. This was greater than the UK average of 29.8PP. Within Wales, the Local Authorities of Blaenau Gwent (47PP) and North Port Talbot (45PP) are among the areas with the largest disability employment gap in the UK (Department for Work and Pensions, 2023).

As of 2021, more working age disabled people had no qualifications than non-disabled people (13% vs. 5%) (Department for Work and Pensions, 2023). The employment patterns of disabled workers in Wales display a greater dependence on part-time work (working 30 hours or less per week) (Department for Work and Pensions, 2023).

Disabled people aged 21 to 64 in Wales are more likely to have no qualifications compared to non-disabled people (13% vs. 7%). The disability employment gap is greater for those disabled people without any qualifications. Additionally, the employment patterns of workers with a disability indicate a greater dependence on part-time work when compared to non-disabled workers in the same age group (41% vs. 29%) (Department for Work and Pensions, 2022b).

### 12.1 Legislation targeting the disabled workforce

The UK and Welsh governments are both working on reducing the disability employment gap. The UK Government is responsible for providing employment support and social security, while the provision of employability and skills assistance is a devolved matter (Senedd Insight, 2023). In the UK, 38,620 people receive Access to Work payments (2% increase from 2021). The majority of those receiving Access to Work payments were Support Workers (n=17,610; 46% of all payments) (Department for Work and Pensions, 2022a).

Devolved initiatives targeting a reduction in the disability employment gap in Wales include:

Targeted Welsh Government policy such as the Employability Plan 2018 (Welsh Government, 2018a).

- The Employability Plan 2018 stated an explicit aim to work with employers to create workplaces that were inclusive and supportive of disabled recruitment and employment.
- The Disability Equality Forum advised on the Employability Plan 2018 and continue to offer advice and guidance to Welsh Government on issues concerning disabled people.
The creation of Disabled People’s Employment Champions as part of the Welsh Governments ‘Right to Independent Living Framework’ (Welsh Government, 2019a).

- Disabled People’s Employment Champions are dedicated roles that actively work with employers in Wales to highlight the benefits of employing disabled people and those with limiting health conditions.
- Employment champions operate within the Social Model of Disability that recognises barriers in society can act to disable people with impairments or health conditions.
- Champions aim to promote the removal of societal barriers that impede participation and progression of employment for disabled people.
The production of the ‘Stronger, fairer, greener Wales: a plan for employability and skills’ (Welsh Government, 2018b).

- The plan acknowledges disparities in disabled employment and commits to achieve improved employment outcomes for the disabled labour force.
- Educational inequalities of disabled people are targeted and opportunities for training and skill development will be widened to allow greater uptake of education in this previously underserved group.

UK Government legislation concerning disability and employment includes the 2010 Equality Act, prohibiting discrimination of disabled individuals in recruitment and employment. The 2010 Equality Act covers Wales and the implications discussed below legally must be adhered to in Wales (UK Legislation, 2010).

### 12.2 Reasonable adjustments/job accommodations

Research suggests that accommodation provided by employers for employees living with mental health disorders helps employees carry out their roles at minimal cost to employers and alleviates the severity of certain mental health disorders (Zafar et al., 2019).

Included under the 2010 Equality Act is the need for reasonable adjustments to be made by employers if an employee requires them to conduct their working responsibilities. Reasonable adjustments defined by the Equality Act (2010) can include, but are not limited to, changes to recruitment processes to ensure fair and equal competition between disabled and non-disabled applicants, making physical changes to the workplace, changes to equipment and flexible hours or working location (UK Legislation, 2010).

Benefits derived from making reasonable adjustments to retain an employee with newly developed or existing needs often offset the costs incurred in making those adjustments. The average cost of replacing a single lost colleague is reported to be around £30,000 (Floresco, 2023). The Department of Work and Pensions (DWP) report the costs of making reasonable adjustments to accommodate disabled colleagues are often low (Department for Work and Pensions, 2022b). The DWP outline other benefits organisations can receive by making reasonable adjustments:

- Enabling disabled employees to continue to work diversifies the workforce and better reflects the diverse range of customers and the community they serve
- Disabled people tend to stay in a job for a longer period than non-disabled colleagues and lower rates of absenteeism are also reported (Department for Work and Pensions, 2022b)

It should be noted that reasonable adjustments relate not only to employment but also to education. Schools and education authorities have had a duty to provide reasonable adjustments for disabled pupils since 2002: originally, under the Disability Discrimination Act 1995 (UK Legislation, 1995); and, from October 2010, under the Equality Act 2010. These adjustments are made to support children and young people to fully participate in education and provide them with the best possible opportunity for educational attainment and career progression.

20% of disabled young people aged 16 to 18 were NEET over a three-year period to September 2021, which rises to 40% for those aged 19 to 24. This compares to 7% and 9%, respectively, for non-disabled young people (Amendola, 2022).

### 12.3 Neurodiversity in the workforce

Severe or specific learning difficulties and autism are the disabilities with the lowest employment rates of 26% and 29%, respectively, in the UK (Department for Work and Pensions, 2023). The Autism All Party Parliamentary Group (APPGA) is a cross-party group made up of Members of Parliament and the House Lords representatives who work together to further the agenda for autism awareness in government. In 2009, the Autism Act was passed that held government accountable to implement a strategy for improving services for autistic adults (National Autistic Society, 2023). A retrospective review of the Autism Act 2009 10 years on by the APPGA identified contributing factors to the persistently low rates of employment amongst working age adults living with autism since the Act was passed. Factors included:

- Barriers to accessing tailored autism support both at the recruitment stage and whilst employed
- Employer awareness and biases towards autism (31% of employers said autistic employees would require too much support; National Autistic Society, 2019)
- The COVID-19 pandemic had significant impacts on employment of people living with autism, which has always been challenging (National Autistic Society, 2023; UK Legislation, 2009).

In Wales, Awtistiaeth Cymru/Autism Wales offers support and guidance to people with autism, carers/parents of someone with autism, individuals seeking autistic people find employment and employers. A comprehensive series of resources can be found at autismwales.org.

## 13 The Welsh language and work

The Welsh language is spoken by 18% of the population of Wales (Welsh Government, 2022d). Between 2011 and 2021, there has been an increase in the number and percentage of the population aged 16 years or older in employment who are able to speak Welsh (from 227,760 [16.6%] in 2011 to 231,400 [16.9%] in 2021) (Welsh Government, 2023p). The reason for this increase is likely because of the Welsh Language (Wales) Measure of 2011; the Welsh Government policy to increase the number of Welsh speakers to 1 million by 2050 (Welsh Government, 2017) and the Welsh Language Commissioners Working Welsh initiatives (Iaith gwaith) (Welsh Language Commissioner, 2023).

In the 2021 Census, 21% of Welsh speakers worked in Professional Occupations, and 12% worked as Process, Plant, or Machine Operatives (Welsh Government, 2023p).

Although it is seen as a positive skill to be able to speak Welsh in Wales, workers from over the Welsh border may not wish to learn the Welsh language, and this could be a potential barrier for working aged individuals who would otherwise consider migrating over the border for employment in Wales (Welsh Language Commissioner, 2023).

**Image 1:**
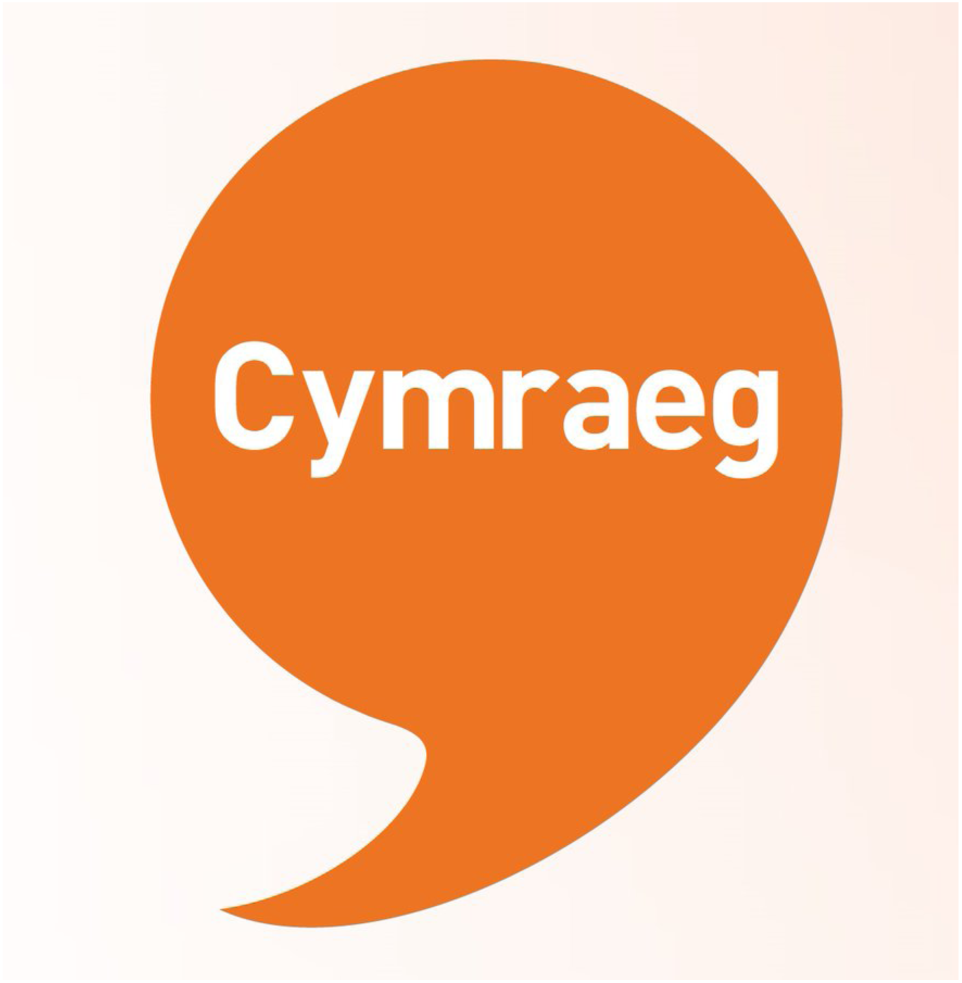
A label signifying a Welsh speaker in the workplace (Cymraeg is the Welsh language). Source: Iaith Gwaith/Working Welsh (Welsh Language Commissioner, 2023)

## 14 Sustainable work practices

### 14.1 Cycle to work scheme

The Welsh Government declared the climate emergency in 2019 and launched ‘Team Wales’. Team Wales is a collaborative approach for public bodies to work together to reach the collective goal of net zero emissions (Welsh Government, 2019c, 2020b). A green economy is an economy that aims to reduce environmental risks and ecological scarcities, and that aims for sustainable development without degrading the environment. It is closely related with ecological economics but has a more politically applied focus (Welsh Government, 2018b).

The ‘Cycle to Work Scheme’ is an initiative that Welsh Government launched to promote healthier lifestyles for employees (Senedd Cymru/Welsh Parliament, 2020). This initiative encourages employees to make cycling their primary means of transportation to reduce carbon emissions (Welsh Government, 2023n). Employees benefit from savings and tax breaks up to 42% off the cost of purchasing their bicycles. The Welsh Government e-MOVE scheme is currently running in five locations across Wales (Rhyl, Swansea, Newtown, Aberystwyth, and Barry). The scheme has saved 600kg of CO_2_ and service users have reported a 39% decrease in car journeys and a 76% positive impact on well-being since the start of the scheme in 2021 (The Bike Shop Wales, 2023).

### 14.2 Smart and flexible working

Retaining staff that have been trained is an important objective of well-being workplace initiatives (Welsh Government, 2023l). A smarter working initiative was launched by Welsh Government, encouraging Welsh businesses to support employees via remote working. This initiative supports vulnerable populations such as disabled employees and employees with caring or childcare responsibilities (Welsh Government, 2022b).

#### 14.2.1 Working from home

Decreased mental well-being and increased loneliness affected more than 45% of the working from home (WFH) population in Wales between November 2020 and January 2021 during the second lockdown of the COVID-19 pandemic (O’Connor et al., 2021). The WFH populations most likely to be affected were workers in their 30s, women, workers living alone, workers with lower mental well-being and workers with pre-existing health conditions. Of those surveyed, 1 in 5 workers reported complete aversion to WFH; conversely, 3 in 5 workers wanted to spend some or all their work week WFH. WFH is not a viable option for all workers, especially workers living in deprived areas, temporarily employed staff, and workers with pre-existing health conditions (Griffiths et al., 2022).

## 15 Barriers to productivity and well-being in the workforce

### 15.1 Gambling and employment

Problem gambling can have a detrimental impact on work and employment outcomes. Gambling is linked to a higher risk of future unemployment (Muggleton et al., 2021). The impacts of gambling include absenteeism, an inability to work, unemployment and reduced performance at work or education (Langham et al., 2016). The financial cost to the UK Government associated with problem gambling is estimated to have cost £77 million in unemployment benefits between 2019 and 2020 (Office for Health Improvement & Disparities, 2023).

### 15.2 Financial wellness

With rising inflation and the current cost of living crisis affecting the UK, many employees are experiencing poor mental health due to worrying about their finances. 60% of people in Wales agreed that rising costs of living negatively affected their quality of life (25% strongly agreed). 87% reported ‘worrying’ about the cost of living, with 38% reporting ‘worrying a lot’ (Public Health Wales, 2023b). People with poor mental health are more likely to experience subsequent reductions in income. The negative impacts on mental health and well-being induced by the cost of living crisis could lead to future financial and health problems, creating an entrenched downward spiral that can persist even if economic conditions improve (Thomson et al., 2022)

Financial issues are the primary stressor to employees (Jaggar & Navlakhi, 2021). Employers are increasingly aware of such issues and are moving to offer tailored financial wellness support. For example, Bangor University offers several non-monetary, financial support interventions for employees. Debt and Financial Stress Support is an intervention that offers guidance and emotional support in managing financial problems. Additionally, workshops are offered about financial awareness and resilience, retirement and pension planning, and will writing services (Bangor University, 2023).

## 16 Review methodology

Stakeholders from the Welsh Government and Public Health Wales asked the BIHMR Rapid Review team to investigate the agreed review question to inform future Welsh Government policy and spending.

**Review Question:** What cost-effective interventions are there to support people in work and assist people to return to the workforce?

Searches were conducted on 2^nd^ June 2023 to identify economic evidence of workforce health and well-being interventions and interventions that encourage employee return to work. To be eligible for inclusion, primary studies or review papers needed to report on economic evaluations, return on investment analyses, costing analyses, or work-related outcomes of economic interest (i.e., employment rates, sickness absence, presenteeism, work productivity). The Population, Intervention, Control and Outcomes (PICO) keywords can be found in Appendix 1. We included evidence from the Organisation for Economic Co-operation and Development (OECD) country settings published between 2017 and 2023. Papers were excluded if full text publications were not available in English or Welsh.

Searches were performed in MEDLINE, Embase and the Cochrane Library. The search strategy performed in MEDLINE can be viewed in Appendix 2. The searches were purposely broad to capture a wide range of different workplace interventions. The search strategy included the terms ‘workplace’, ‘wellness at work’, ‘healthy workplace’ and health economics terms to capture economic evaluations and costing studies such as ‘cost-effectiveness’, ‘cost-benefit’ and ‘cost analyses’.

The review identified a wide range of workplace well-being interventions targeting several different areas, including but not limited to, mental health, substance abuse, smoking, weight management, physical activity, and interventions for specific populations such as health care workers (HCWs). Examples of published economic evidence of different workplace health and well-being interventions supporting different subgroups identified from the searches are presented below. The Preferred Reporting Items for Systematic reviews and Meta-Analyses (PRISMA) flow diagram can be found in Appendix 3 and the data extraction tables can be found in Appendix 4. The final number of eligible papers was 76. Summary statistics regarding the Welsh workforce can be found in Appendix 5.

## 17 Economic evidence for interventions supporting well-being to enable retention and return to the workforce

### 17.1 Common mental health conditions

The costs of mental health in Wales are estimated at £7.2 billion per year and from 2019 to 2020 £810 million was spent on mental health problems (Welsh Government, 2023i). Common mental health problems such as anxiety, depression and unmanageable stress affect one in six employees in Wales each year (Belloni et al., 2022). The COVID-19 pandemic had a negative impact on population general health with specific groups of people affected which included: those with pre-existing mental health concerns, NHS/social care workers and key workers, low-income earners, and the self-employed (Parliament, 2020).

Common mental health problems often affect the workplace environment, significantly impacting both employees and employers. Conditions such as anxiety and depression may lead to reduced productivity, increased absenteeism, and strained interpersonal dynamics among co-workers (Ling, 2023). Additionally, workplace stressors, such as long hours, lack of work-life balance, and limited support from management, can contribute to the development or worsening of mental health challenges. Creating a mentally healthy work environment involves fostering open communication, promoting psychological well-being, and offering access to resources such as therapy or counselling (Ling, 2023). Prioritising employee mental health enhances job satisfaction, retention, and contributes to a more productive and compassionate work culture (Brunges & Foley-Brinza, 2014).

Tele coaching had positive effects on mental health outcomes in the workplace during the COVID-19 pandemic (Barak et al., 2008; Sharma & Bhaskar, 2020). Tele coaching is coaching at a distance using telecommunications. It can be used to improve mental health along with supporting self-care. Tele coaching has emerged as a valuable tool with positive effects on mental health outcomes in the workplace. As remote work has become the new norm, individuals face heightened feelings of isolation, stress, and uncertainty. Tele coaching, through its virtual platforms, has provided a convenient and accessible means for employees to receive personalised support and guidance. Studies have indicated that remote coaching interventions can lead to improved mental well-being and stress reduction (Barak et al., 2008). Professionals offering tele coaching help individuals develop coping strategies, manage work-related stressors, and navigate the blurred boundaries between work and home life. This form of remote coaching also encourages self-care practices and resilience-building, contributing to improved mental well-being. By tailoring sessions to address individual concerns and fostering a sense of connection, tele coaching has played a significant role in mitigating the negative impacts of the pandemic on mental health within the workplace setting.

Mental health problems have an adverse effect on the ability of people to work, costing the Welsh Government over £1.2 billion a year including state benefits costs, lost tax and National Insurance revenue, and NHS costs (Farmer & Stevenson, 2017). The cost of mental health problems at work to the Welsh economy may be much higher with between £3.5 billion (Friedli & Parsonage, 2009) and £4.7 billion every year lost in terms of lost output, costs to employers and NHS costs. The full societal costs of poor mental health in Wales could be as high as £9.5 billion when including the costs to health and social care (£1.4 billion) and considering the high human cost of mental health problems (£4.6 billion). The UK public health guidance on ‘Mental well-being at work’ states that organisation-wide approaches to promoting mental well-being can produce important net economic benefit and that performing annual audits of employee well-being would produce financial gains; of the order of £100 million per annum (National Institute for Health and Care Excellence, 2022b). Workplace mental health interventions can offer a positive ROI with up to £9 generated for every £1 spent (Deloitte, 2022; Knapp et al., 2011). There are many examples of interventions that promote mental health and well-being at work in the literature. These examples highlight the cost-effectiveness of programmes to improve mental health in the workplace. Tufts Be Well at Work is a telephone-based programme designed to aid the workplace functioning of employees living with depression in the United States of America (USA). The programme is an effective intervention for improving productivity, especially when compared with conventional interventions. It also has low running costs and a positive ROI (in the three randomised controlled trials (RCTs), the ROI was 5.4:1, 5.2:1, and 1.6:1) (Lerner et al., 2021). Senedd Insight, a Welsh Government consultant agency, hosted the ‘Creating Mentally Healthy Workplaces Wales Conference’ in October 2023 to promote prevention, healthy workplace practice, cultivation of healthier environments, and effective means of leadership and crisis management (Senedd Insight, 2023). RESPECT is a four-hour face-to-face mental health training programme for managers which aims to better understand employee mental health in Australia. This programme led to a reduction in workplace absenteeism and an ROI of £9.98 for every £1 spent on manager training (Milligan-Saville et al., 2017). Individual psychological support for health care workers in Italy dealing with occupational distress yielded an ROI of €2.73 for every €1 invested (Dalmasso et al., 2021). Problem-solving based intervention (PSI) for common mental health disorders reduced the socio-economic burden when compared with care as usual in Sweden. PSI was not cost-effective for the employer, but it was cost-beneficial to society (Van De Poll et al., 2020).

Early intervention in the workplace for common mental health disorders and targeted effective treatment for at-risk employees can be cost saving for businesses and the NHS (Friedli & Parsonage, 2009). Organisation-wide primary prevention initiatives may offer a greater ROI than ‘reactive’ intervention at a later stage (e.g., secondary, or tertiary prevention) with culture change, or awareness raising workplace health promotion interventions offering around £8 ROI for every £1 spent, compared with targeted psycho-social mental health treatments for depression generating up to £5 for every £1 investment (Deloitte, 2022). There is evidence from the NHS in Wales that interventions such as yoga can be cost-effective in terms of reducing absenteeism due to musculoskeletal disorders. For every £1 spent on yoga, there is an estimated £10.17 societal benefit generated due to increased productivity at work (Public Health England, 2019). Embedding economic evaluations into future research on workplace health outcomes would help enable the identification of cost-effective workplace health programmes (Brunton et al., 2016). The management of fit/sick notes should also be reviewed in a future study (Public Health Wales, 2023a). See Table 2 for recent included studies relating to common mental health disorders.

**Table 2.** Economic studies relating to common mental health disorders.

| Author | Date and Country | Aim | Intervention | Length of follow-up | Type of study | Population | Findings | Type of economic analysis and main finding | *Quality rating |
| --- | --- | --- | --- | --- | --- | --- | --- | --- | --- |
| **Lerner et al.** | 2021<br>USA | To test whether the Veterans Health Administration (VHA) integrated care (IC) program combined with telephonic work-focused counselling, known as Be Well at Work (BWA), is superior to IC alone. | The BWA intervention was based on stress and coping theory in which successful mental health and functional outcomes are related to responding effectively to psychosocial challenges. | 4 months | RCT with ROI analysis. | 253 patients (mean [Standard Deviation [SD]] age, 45.7 [11.6] years; 218 [86.2%] men; 135 [53.4%] white). IC control group (n=114); intervention group received IC plus 'Be Well at Work' (n=139). | At the 4-month follow-up, patients who received IC plus BWA had greater reductions in at-work productivity loss (adjusted effect, -1.7; 95% Confidence Interval (CI), -3.1 to -0.4; P = .01) and depression symptom severity (adjusted effect, -2.1; 95% CI, -3.5 to -0.7; P = .003). The improvements from IC plus BWA persisted 4 months after intervention (at-work productivity loss mean difference, -0.5; 95% CI, -1.9 to 0.9; P = .46; depression symptom severity mean difference, 0.6; 95% CI -0.9 to 2.1; P = .44). The cost per patient participating in BWA was \$690.98, and the return on investment was 160%. | ROI.<br>The cost per patient participating in the Be Well at Work programme was \$691 and the ROI was 160%. | Low |
| **Milligan-Saville et al.** | 2017<br>Australia | To investigate the effect of mental health training on managers' knowledge, attitudes, confidence, and behaviour towards employees with mental health problems, and its effect on employee sickness absence. | Manager mental health training within a large Australian fire and rescue service. | 6 months | RCT with ROI. | The intervention group (n=46) received face-to-face RESPECT Manager Training Programme, which combined mental health knowledge and communication training. | During the 6-month follow-up, the mean rate of work-related sick leave decreased by 0.28PP from a pre-training mean of 1.56% (Standard Error (SE) 0.23) in the intervention group and increased by 0.28PP from 0.95% (0.20) in the control group (p=0.049), corresponding to a reduction of 6.45 hour per employee per 6 months. | ROI.<br>A 4-hour manager mental health training programme could lead to a significant reduction in work-related sickness absence, with an associated ROI of £9.98 for £1 spent on such training. | Moderate |
| **Van De Poll et al.** | 2020<br>Sweden | To assess the cost-benefit and cost-effectiveness of a work-directed intervention implemented by the occupational health service (OHS) for employees with common mental disorders (CMD) or stress-related problems at work. | An occupational health workplace intervention. | 12 months | Cost-benefit and cost-effectiveness. | Problem Solving Based Intervention (PSI) group (n= 41).<br>Care as usual group (n=59). | Costs for long-term sickness absence were higher for care as usual, whereas costs for short-term sickness absence and production loss at work were higher for the PSI group. Due to these costs, PSI was not cost-effective from the employer's perspective. However, PSI was cost-beneficial from a societal perspective. | A cost-effectiveness analysis showed that a one-day reduction of long-term sickness absence costed on average €101 for the PSI group, a cost that primarily was borne by the employer. | High |

\*The Joanna Briggs Institute (JBI) tool used to rate the quality of the studies is designed for full economic evaluations and may have disadvantaged the quality rating of partial economic evaluations such as return on investment analyses.

### 17.2 Workers with rheumatoid arthritis

The evidence on the relative cost-effectiveness of workplace interventions to support workers with rheumatoid arthritis is mixed. A cost-effectiveness analysis and cost-utility analysis of a workplace intervention to improve productivity among workers with rheumatoid arthritis did not produce positive economic outcomes (Noben et al., 2017). Findings indicated that the intervention was more costly and less effective when compared to the care as usual group (Noben et al., 2017).

### 17.3 Musculoskeletal disorders

Musculoskeletal conditions such as lower back pain, neck pain, shoulder pain, leg pain, foot pain and elbow pain reduce levels of paid productivity (Jones et al., 2019). A cost-benefit analysis of a participatory return to work intervention for temporary agency and unemployed workers sick-listed due to musculoskeletal disorders in the Netherlands indicated that although the programme was more effective and had the potential to achieve a sustainable contribution of vulnerable workers to the labour force, it was also more costly than usual care (Vermeulen et al., 2013).

### 17.4 Severe mental health conditions

The number of people in Wales living with severe mental health disorders rose from 12% before the COVID-19 pandemic to 28% by April 2020 (Wales Fiscal Analysis, 2021). The total cost of poor mental health has increased from £42-45 billion (2019 pre-pandemic) to £53-56 billion (2020-2021 during and post-pandemic) an increase of 25%. These costs include absenteeism, presenteeism and staff turnover. The annual costs per employee in Wales are among the highest as a percentage in the UK at 68% of earnings (£1,768 per employee per year). Early proactive interventions such as culture change and prevention that span the entire organisation yield an ROI of £5.30 for every £1 invested (Deloitte, 2022). Increasing evidence suggests that employer intervention can improve mental health via means such as risk and stress management, good working conditions, encouragement of autonomy, supporting work-life balance, a clear path for progression, and the absence of harassment and bullying (McDaid & Park, 2022; National Institute for Health and Care Excellence, 2022b).

The employment rate for people with severe mental health illness (including schizophrenia and mood disorders) is significantly lower than both the general population and people living with a disability, including those with common mental health conditions (Booth et al., 2014). In terms of ROI, the benefits gained from employment include personal health and social benefits, with less reliance on social care translating into savings. An ROI of £1.04 for every £1 spent has been found for placement support programmes in the North West and Yorkshire and Humber regions of England (Booth et al., 2014).

A pilot ROI analysis evaluating a workplace disability management programme for HCWs in an Italian paediatric hospital found significant returns in terms of productivity and reduced absenteeism rates. The programme assessed worker disabilities and provided tailored reasonable adjustments to the work environment or work roles. The total intervention programme cost €14,931 and provided a ROI of €28 per €1 invested, evidencing significant benefit. Significant falls in absenteeism (days absent) were observed at 6-month follow-up (23 to 6 days) and 12-month follow-up (41 to 15 days) (Camisa et al., 2020). The most common reasonable adjustments were internal reorganisation of work duties (relieving HCWs of duties too heavy for their pathology), proposal to company management for environmental, ergonomic, structural, and technological improvement measures, paths of psychosocial support and/or health promotion interventions, and a change of working destination or change of job. The three categories of work disability in the study were work and extra-work discomfort (mainly of a psychological nature linked to work-related stress), musculoskeletal pathology due to biomechanical overload and other problems related to the presence of serious pathologies (especially neoplasms) (Camisa et al., 2020).

Two economic evaluation studies have assessed the cost-effectiveness of interventions for individuals with traumatic brain injury and veterans with spinal cord injury (Radford et al., 2018; Sutton et al., 2020). In the USA, a cost-utility analysis of supported employment for veterans with spinal cord injury was found to be more effective but not cost-effective when compared to usual care (Sutton et al., 2020). In England, a feasibility RCT demonstrated favourable findings in terms of an early-stage cost-utility analysis of specialist vocational rehabilitation to increase return to work among individuals following traumatic brain injury (Radford et al., 2018). See Table 3 for studies relating to severe mental health disorders.

**Table 3.**
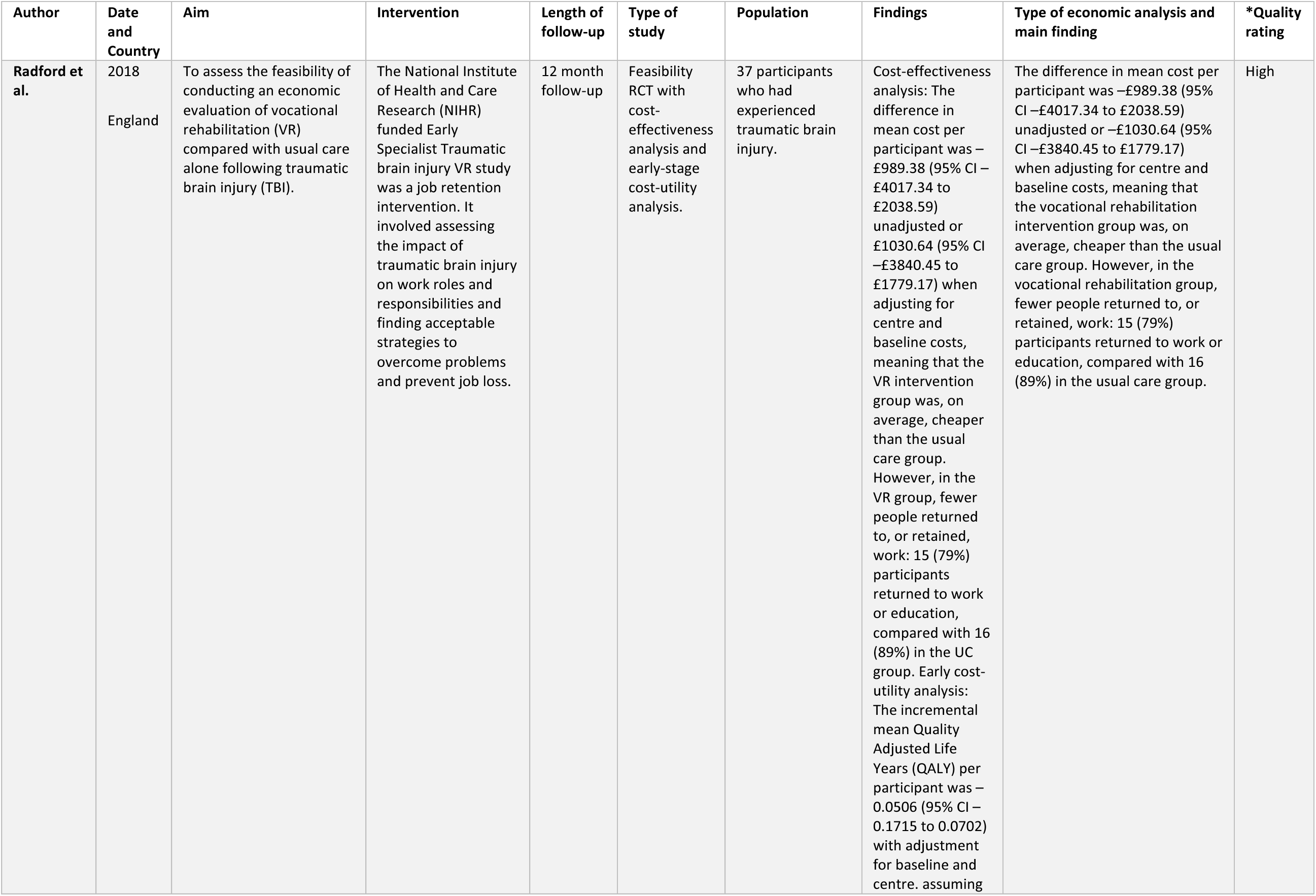
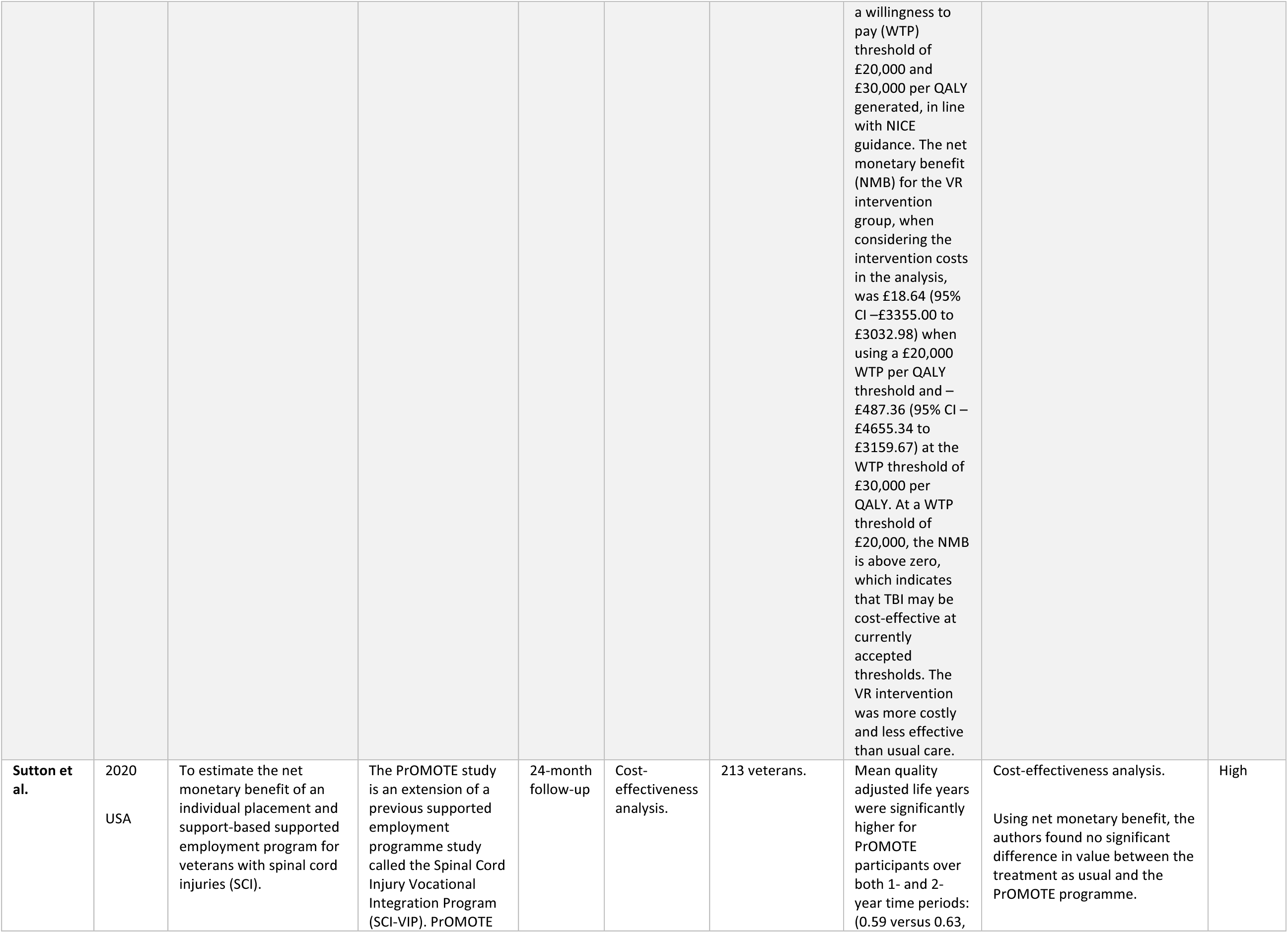
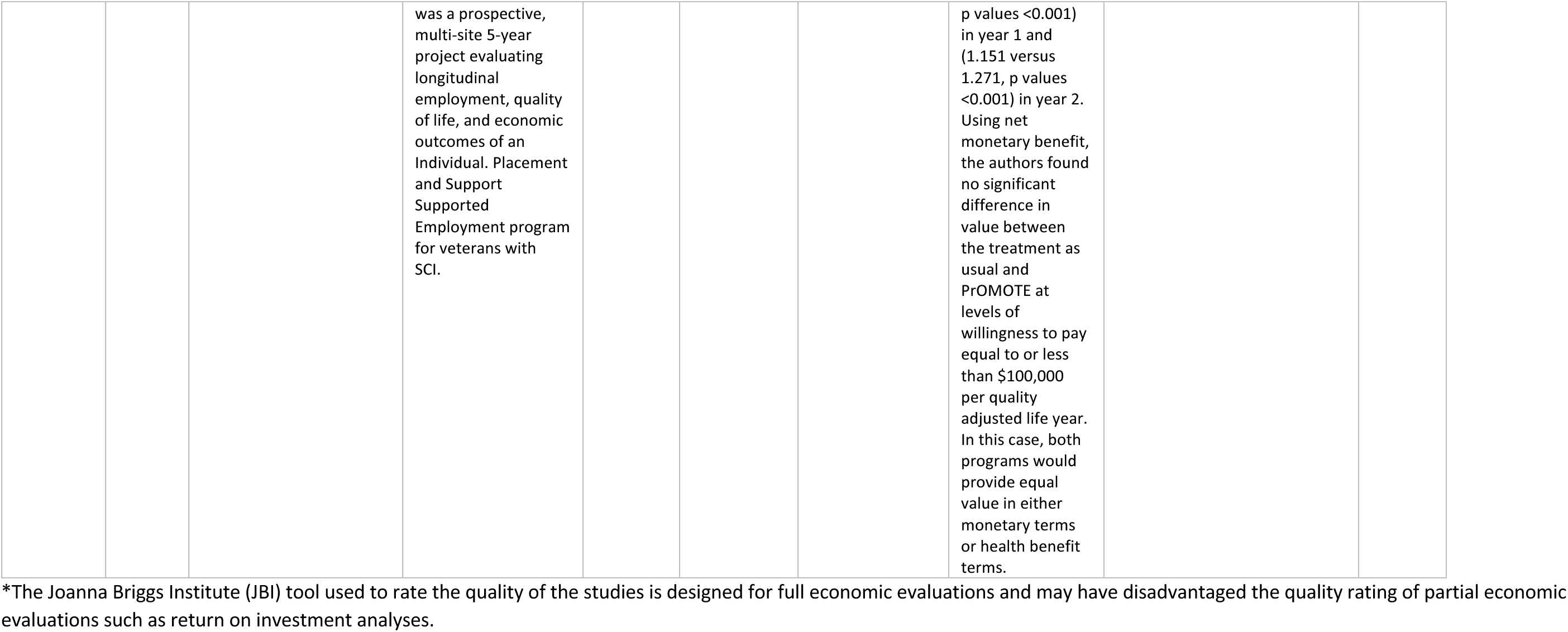
Economic studies relating to severe mental health disorders.

| Author | Date and Country | Aim | Intervention | Length of follow-up | Type of study | Population | Findings | Type of economic analysis and main finding | *Quality rating |
| --- | --- | --- | --- | --- | --- | --- | --- | --- | --- |
| Radford et al. | 2018<br>England | To assess the feasibility of conducting an economic evaluation of vocational rehabilitation (VR) compared with usual care alone following traumatic brain injury (TBI). | The National Institute of Health and Care Research (NIHR) funded Early Specialist Traumatic brain injury VR study was a job retention intervention. It involved assessing the impact of traumatic brain injury on work roles and responsibilities and finding acceptable strategies to overcome problems and prevent job loss. | 12 month follow-up | Feasibility RCT with cost-effectiveness analysis and early-stage cost-utility analysis. | 37 participants who had experienced traumatic brain injury. | Cost-effectiveness analysis: The difference in mean cost per participant was –£989.38 (95% CI –£4017.34 to £2038.59) unadjusted or –£1030.64 (95% CI –£3840.45 to £1779.17) when adjusting for centre and baseline costs, meaning that the VR intervention group was, on average, cheaper than the usual care group. However, in the VR group, fewer people returned to, or retained, work: 15 (79%) participants returned to work or education, compared with 16 (89%) in the UC group. Early cost-utility analysis: The incremental mean Quality Adjusted Life Years (QALY) per participant was –0.0506 (95% CI –0.1715 to 0.0702) with adjustment for baseline and centre. assuming<br>a willingness to pay (WTP) threshold of £20,000 and £30,000 per QALY generated, in line with NICE guidance. The net monetary benefit (NMB) for the VR intervention group, when considering the intervention costs in the analysis, was £18.64 (95% CI –£3355.00 to £3032.98) when using a £20,000 WTP per QALY threshold and –£487.36 (95% CI –£4655.34 to £3159.67) at the WTP threshold of £30,000 per QALY. At a WTP threshold of £20,000, the NMB is above zero, which indicates that TBI may be cost-effective at currently accepted thresholds. The VR intervention was more costly and less effective than usual care. | The difference in mean cost per participant was –£989.38 (95% CI –£4017.34 to £2038.59) unadjusted or –£1030.64 (95% CI –£3840.45 to £1779.17) when adjusting for centre and baseline costs, meaning that the vocational rehabilitation intervention group was, on average, cheaper than the usual care group. However, in the vocational rehabilitation group, fewer people returned to, or retained, work: 15 (79%) participants returned to work or education, compared with 16 (89%) in the usual care group. | High |
| **Sutton et al.** | 2020<br>USA | To estimate the net monetary benefit of an individual placement and support-based supported employment program for veterans with spinal cord injuries (SCI). | The PrOMOTE study is an extension of a previous supported employment programme study called the Spinal Cord Injury Vocational Integration Program (SCI-VIP). PrOMOTE was a prospective, multi-site 5-year project evaluating longitudinal employment, quality of life, and economic outcomes of an Individual. Placement and Support Supported Employment program for veterans with SCI. | 24-month follow-up | Cost-effectiveness analysis. | 213 veterans. | Mean quality adjusted life years were significantly higher for PrOMOTE participants over both 1- and 2-year time periods: (0.59 versus 0.63, p values <0.001) in year 1 and (1.151 versus 1.271, p values <0.001) in year 2. Using net monetary benefit, the authors found no significant difference in value between the treatment as usual and PrOMOTE at levels of willingness to pay equal to or less than \$100,000 per quality adjusted life year. In this case, both programs would provide equal value in either monetary terms or health benefit terms. | Cost-effectiveness analysis.<br>Using net monetary benefit, the authors found no significant difference in value between the treatment as usual and the PrOMOTE programme. | High |

\*The Joanna Briggs Institute (JBI) tool used to rate the quality of the studies is designed for full economic evaluations and may have disadvantaged the quality rating of partial economic evaluations such as return on investment analyses.

### 17.5 Vaccinations

With respect to common ailments such as influenza, vaccinating the workforce against certain illnesses has been shown to be cost-effective for health and social care workers, but evidence of cost-effectiveness across the general workforce is less convincing. This is because the ripple effects are less than in a health care setting and immunising an employee is mainly only beneficial to immediate family and not others within the community (Edwards et al., 2019). Identified economic analyses assessing vaccinations in the workforce concerned influenza and COVID-19 vaccinations only. The evidence is presented below.

#### 17.5.1 Influenza (flu)

A cost-benefit analysis conducted during 2017 and 2018 was part of an influenza vaccine observational study (Ferro et al., 2020). For every participant data were collected on seasonal influenza immunisation status and sick-leave days. Sick-leave days were compared among the influenza epidemic period and the previous one between vaccinated and unvaccinated and any difference in days of absence was caused by seasonal influenza. The monthly mean cost for sickness absences per employee was significantly higher for an unvaccinated individual compared to one vaccinated, respectively €129.00 and €54.00 (p = 0.028). The overall net saving estimated was €314.00 per person vaccinated (Ferro et al., 2020).

A European modelling study of workplace influenza vaccination found that 90% of vaccination coverage could reduce the burden of influenza both on the workforce and the wider population. The average cost of a vaccine was €10 per vaccinated employee (Verelst et al., 2021).

Cost–benefit analysis was the most common economic evaluation method for interventions against influenza at the workplace (Ofori et al., 2022). All except two cost–benefit analyses on vaccination indicated cost savings or cost–effectiveness, and only one study was identified to have assessed nonpharmacological interventions. The average net benefit of vaccination per employee was $658.28 based on a study from Spain (Fernández, 2006). For those studies reporting their results as the cost–benefit ratio, a study from the USA reported a cost–benefit ratio of 0.1219 meaning that costs outweighed benefits (Akazawa et al., 2003) and a study from Brazil reported a cost-benefit ratio of 1:2.47 (Burckel et al., 1999). A net societal cost of $40.89 per vaccinated person compared with no vaccination was reported in a study from the USA (Bridges et al., 2000). See Table 4 for studies relating to flu vaccinations.

**Table 4.** Studies relating to influenza vaccinations.

| Author | Date and Country | Aim | Intervention | Length of follow-up | Type of study | Population | Findings | Type of economic analysis and main finding | *Quality rating |
| --- | --- | --- | --- | --- | --- | --- | --- | --- | --- |
| **Ferro et al.** | 2020<br>Italy | To analyse the extent of absences due to illness following a voluntary influenza immunisation program among the Komatsu Italia Manufacturing company's personnel during the flu season 2017-2018. | Influenza vaccine 2017-2018. | 12 months | Cost-benefit analysis. | Unvaccinated employees over the influenza season 2017-2018. | During the influenza period, the monthly mean of sick-leave days per employee was significantly lower among the vaccinated than the unvaccinated. | The monthly mean cost for sickness absences per employee was significantly higher for an unvaccinated employee compared to one vaccinated, respectively €129.00 and €54.00 ($p = 0.028$). The overall net saving estimated was €314.00 per person vaccinated. | High |
| **Verelst et al.** | 2021<br>UK, Belgium & the Netherlands | To estimate the cost benefits of employer funded flu vaccination in Belgium with a dynamic transmission model. | Workplace influenza vaccination. | Unclear (this was a modelling study). | Economic modelling study. | Modelled using Belgium population data. | Workplace vaccination reduced the burden of influenza both on the workplace and in the population at large. | Compared to the current vaccine coverage – 21% in the population at large – an employee vaccine coverage of 90% could avert an additional 355,000 cases, of which about 150,000 in the employed population and 205,000 in the unemployed population. While seasonal influenza vaccination has been cost saving, on average at about €10 per vaccinated employee, the cost-benefit analysis was prone to between-season variability. | High |

\*The Joanna Briggs Institute (JBI ) tool used to rate the quality of the studies is designed for full economic evaluations and may have disadvantaged the quality rating of partial economic evaluations such as return on investment analyses.

#### 17.5.2 COVID-19 and Long-COVID

In January 2020, the COVID-19 virus was identified and quickly spread around most of the world, and since then, COVID-19 has had a significant impact on the way that people work, with many more working from home than in the years preceding the COVID-19 pandemic (Hupkau & Petrongolo, 2020). One in four employees now have a hybrid working week, and 16% of UK workers work exclusively from home in 2023. Up to 69% of professional occupations, such as accountants and lawyers, are most likely to be working remotely (Office for National Statistics, 2023b). During the COVID-19 lockdowns, the Coronavirus Job Retention Scheme (CJRS) (more widely known as furlough) was launched by the UK Government in March 2020 to support businesses to pay their employees. The scheme supported 1.3 million businesses and 11.6 million jobs between March 2020 and September 2021. Most employees were working in the hospitality, construction, and recreation industries. The cost of the CJRS scheme was £70 billion (Francis-Devine et al., 2021). During this time, the government also spent £88 billion on developing a vaccine to reduce the spread of COVID-19 (Health in developing countries, 2022).

There have been many individuals infected with COVID-19 left with poorer quality of life due to Long-COVID. Long-COVID refers to signs and/or symptoms that develop following COVID-19 infection which continue for more than 12 weeks and are not explained by an alternative diagnosis (World Health Organisation (WHO), 2022). Some of the common symptoms include fatigue, shortness of breath, chest pain, and neurocognitive impairment, which often overlap and fluctuate in severity. Some of the symptom’s impact on function and could impede returning to work after the initial infection (Madan et al., 2021). Working-aged people are less likely to participate in the labour market after developing Long-COVID symptoms than they were before being infected with COVID-19. This relationship between self-reported Long-COVID and inactivity for reasons other than education or retirement is strongest among people aged over 50 years (Ayoubkhani, 2022).

#### 17.5.3 Gender

Men were more likely to die from COVID-19 than women (Capuano et al., 2020). There was an 18% difference in the total number of COVID-19-related deaths for men (63,700) and women (53,300) between March 2020 and January 2021 in England and Wales (Office for National Statistics, 2022). Reasons for this may include immune system activity and its modulation by sex hormones, coagulation pattern, and pre-existing cardiovascular diseases as well as effects from smoking and drinking habits (Capuano et al., 2020).

#### 17.5.4 Minority groups

COVID-19 disproportionately affected individuals from ethnic minority communities in the UK. During the first wave of the COVID-19 pandemic (24 January 2020 to 11 September 2020), people from all ethnic minority groups (except for women in the Chinese or “White Other” ethnic groups) had higher rates of death involving the coronavirus compared with the White British population. The rate of death involving COVID-19 was highest for the Black African group (3.7 times greater than for the White British group for men, and 2.6 greater for women), followed by the Bangladeshi (3.0 for men, 1.9 for women), Black Caribbean (2.7 for men, 1.8 for women) and Pakistani (2.2 for men, 2.0 for women) ethnic groups (Office for National Statistics, 2021e).

### 17.6 Alcohol misuse

Alcohol misuse or excessive alcohol consumption puts individuals at increased risk with adverse health and social consequences (Glesson et al., 2019). Lost productivity due to alcohol misuse costs the UK economy more than £7 billion annually, and an estimated 167,000 working years are lost to alcohol consumption every year (Public Health England, 2016). The effects of alcohol consumption are long lasting, and therefore work productivity can be affected in the long and short term. Workers may attend work hungover or still under the influence of alcohol from the night before, consume alcohol before work, or during the working day (Alcohol Change UK, 2018). Heavy alcohol consumption also leads to more serious health issues, such as alcohol-related liver disease (ALD) and other cardiovascular complications including heart failure; gout, and cancer, injuries, brain disease like dementias and Wernicke-Korsakoff’s encephalopathy (Meza et al., 2022).

A recent systematic review included 39 articles, with 28 of these describing primary research and 11 reviews, most of which focused solely on alcohol use. Heterogeneity between studies concerning intervention and evaluation design limited the degree to which findings could be synthesised. Targeted brief interventions and universal substance use screening were found to be useful in reducing alcohol consumption (Morse et al., 2022). The use of alcohol increases the number of accidents and mistakes because of the reduction in concentration ability. In the USA in 2010, a screening and referral alcohol abuse programme with the workforce was modelled (Quanbeck et al., 2010). The Screening, Brief Intervention, and Referral to Treatment Programme (SBIRT) benefit-cost ratio was found to be 4.4:1, providing convincing evidence that the SBIRT programme could be beneficial for employers in companies where there are workers with problem drinking profiles.

Alcohol-related presenteeism (impaired work performance associated with alcohol consumption) has been identified as an under-researched topic in the research literature (Thørrisen et al., 2019). Evidence supports the view that employee alcohol consumption may be associated with impaired work performance (Jurek & Rorat, 2017; Schou, 2016). Due to low research quality and lack of longitudinal designs, existing evidence should still be characterised as inconclusive regarding the prevalence, nature, and impact of alcohol-related presenteeism in the workforce (Thørrisen et al., 2019).

Those who live and work in cultures where drink culture is the norm are also at more risk of alcohol-related harm (Alcohol Change UK, 2018). Those at most risk of alcohol related harms are:

- Shift workers
- Workers working in poor conditions
- Workers under stress
- Workers with low job security

##### Box 2 Facts about alcohol related harm (Public Health England, 2016)

- 40% of employers mention alcohol as a significant cause of low productivity
- Between 3% and 5% of all work absence is caused by alcohol consumption
- 35% of people report they have noticed colleagues under the influence of drugs and alcohol at work
- 25% of people report that drugs or alcohol have affected them at work, with 23% reporting they had experienced decreased productivity as a result

There have been many interventions to reduce presenteeism due to alcohol abuse worldwide. Interventions include:

- Theoretical based – mental simulation (Hagger et al., 2011)
- Alcohol screening
- SBIRIT (Quanbeck et al., 2010)
- Community based interventions such as the North Wales Alcohol Harm-Reduction Strategy (Betsi Cadwaladr University Health Board, 2020)

The effectiveness of interventions to reduce risky patterns of alcohol consumption is mixed (Hagger et al., 2011; Wolfenden et al., 2018). Despite the limited number of implementations studies, the evidence suggests that interventions that cause low burden, that are confidential, and those which have low psychological resistance lead to stronger intentions to reach the desired outcome (Hagger et al., 2011).

Reviews of the evidence suggest interventions that are likely to be particularly effective in reducing alcohol consumption include: increasing alcohol prices; reducing the availability of outlets selling alcohol and restricting the marketing of alcohol (Jackson & Johnson, 2010; National Institute for Health and Care Excellence, 2022a; World Health Organisation (WHO), 2009).

In Wales, minimum (alcohol) unit pricing (MUP) was based loosely on the Scottish Governments’ model of MUP. On 1^st^ May 2018, Scotland introduced an MUP of 50 pence per alcohol unit. MUP in Scotland was associated with a significant 13.4% reduction (95% CI –18.4 to –8.3; p=0.0004) in deaths wholly attributable to alcohol consumption. There was also a significant decrease in hospitalisations wholly attributable to alcohol consumption (–8.3 to 0.3; p=0.064) (Wyper et al., 2023). Effects were driven by significant improvements in chronic outcomes, particularly alcoholic liver disease. MUP legislation was associated with a reduction in deaths and hospitalisations wholly attributable to alcohol consumption in the four most socioeconomically deprived areas in Scotland (Wyper et al., 2023).

The MUP was introduced in Wales in March 2020, and the law includes a formula for calculating the MUP. This formula is made up of the MUP of 50 pence, the strength and volume of the alcohol. This formula has been deliberately chosen to target high alcohol content drinks, which tend to be consumed by those who are more at risk of harm from alcohol consumption (Welsh Government, 2020a). By October 2020, 10% of drinkers had reduced their alcohol consumption due to the introduction of MUP in Wales (Alcohol Change UK, 2020). There are no up-to-date figures showing the same direction of travel since the COVID-19 pandemic, which led to an increase in alcohol and illicit drug consumption (Holloway et al., 2022). Most drinkers reported little change in either the type or brand of alcohol that they consumed. One of the reasons for the lack of reduction in alcohol consumption was the fact that many people, especially those living in border towns, would travel to England to buy alcohol at lower prices (Buhociu et al., 2021). Drinkers in Wales also buy alcohol directly from breweries in England as England does not currently have MUP.

#### 17.6.1 Gender

Men are more likely to die of alcohol-related factors than women, and this is a consistent pattern across the UK. Rates of deaths amongst men associated with alcohol were twice those of women. With rates ranging between 17.3 and 17.8 deaths per 100,000 men in 2020, compared with rates ranging between 8.0 and 8.6 deaths per 100,000 women (Office for National Statistics, 2021c).

#### 17.6.2 Minority groups

In Wales, people from ethnic minority groups drink less and are more likely to abstain from alcohol than their White counterparts. There are higher rates of harmful drinking among certain groups, for example, older Irish men and men of Sikh religion (Institute of Alcohol Studies, 2020; Public Health England, 2016). There is a lack of evidence around alcohol use and support needs, particularly for ethnic minority women; ethnic minority prisoner populations; refugees and asylum seekers; LGBT individuals belonging to ethnic minority groups, and non-practicing individuals from religious minorities (Glesson et al., 2019).

### 17.7 Illicit drug use and prescription drug abuse and employment

Literature searches identified two economic evaluations and two systematic reviews exploring the cost-effectiveness of interventions targeting substance abuse to improve work-related outcomes (de Oliveira et al., 2020; Morse et al., 2022; Orme et al., 2023; Rognli et al., 2023).

An economic evaluation conducted in the US assessed the cost-effectiveness of substance abstinence contingent wage supplements to prevent substance abuse and increase unemployment rates (Orme et al., 2023). Participation in the abstinence contingent wage supplement programme increased employment rates and negative drug tests among study participants. The incremental cost-effectiveness ratio (ICER) was $1,437 per additional participant with a negative drug test result and $915 per additional person in employment at the end of the 12-month study period (Orme et al., 2023).

In Norway, a cost-effectiveness simulation study based on previously published data was conducted to estimate the costs and outcomes of employment support integrated into substance abuse treatment (Rognli et al., 2023). Three scenarios were compared: treatment as usual (TAU), a self-guide and workshop with TAU, and TAU and individual placement support (IPS). IPS was found to be more effective and cost-effective after 6 months to 2 years in comparison to the two other interventions (Rognli et al., 2023).

A systematic review of workplace mental health and substance use interventions identified moderate strength evidence that cognitive behavioural therapy is cost saving and cost-effective (de Oliveira et al., 2020). The review also identified convincing evidence that occupational therapist interventions to decrease absenteeism and increase return to work are cost saving and cost-effective (de Oliveira et al., 2020).

A second systematic review assessing the cost-effectiveness of workplace interventions for the prevention and treatment of substance abuse presented mixed evidence on the cost-effectiveness from two cost-utility analyses and one ROI study (Morse et al., 2023). See Table 5 for studies relating to drug use.

**Table 5.** Economic studies relating to drug use.

| Author | Date and Country | Aim | Intervention | Length of follow-up | Type of study | Population | Findings | Type of economic analysis and main finding | *Quality rating |
| --- | --- | --- | --- | --- | --- | --- | --- | --- | --- |
| Orme et al. | 2023<br>USA | To estimate the cost-effectiveness of Abstinence-Contingent Wage Supplement (ACWS) compared with usual care. | ACWS compared with usual care. | 12 months. | RCT with cost and cost-effectiveness of abstinence contingent wage supplements. | Intervention group (n=44), control group (n=47). | Two time-points: the percentage of negative drug (opiates and cocaine) tests and the percentage of participants employed at the end of the intervention and post-intervention periods. The ICERs of the ACWS intervention and usual care show the ACWS intervention was economically dominated by usual care. | ACWS cost \$11,310 per participant over the 12-month intervention period. Total intervention and health care costs per participant over the intervention period were \$20,625 for usual care and \$30,686 for ACWS. The ICERs of the ACWS intervention and usual care show the ACWS intervention was economically dominated by usual care. | High |
| Rognli et al. | 2022<br>Norway | To model the potential socioeconomic value of employment support integrated in substance use disorder (SUD) treatment. | (1) treatment as usual (TAU). (2) TAU and a self-help guide and a workshop. (3) TAU and Individual Placement and Support (IPS). | One-year time horizon for the interventions and a 10-year time horizon for the socioeconomic effects of the interventions. | RCT model simulation study with cost-effectiveness. | Individuals having substance use disorder treatment. | Compared to the two alternative interventions, IPS was cost-effective and more beneficial after six months to two years. | Assuming a 40% employment rate and full-time employment (100%) for 10 years following IPS, the authors found a 10-year socioeconomic effect of €18,732,146. The corresponding effect for the more conservative TAU + IPS simulation assuming 40% part-time positions (25%) for five years, was €2,519,906. | High |

\*The Joanna Briggs Institute (JBI) tool used to rate the quality of the studies is designed for full economic evaluations and may have disadvantaged the quality rating of partial economic evaluations such as return on investment analyses.

### 17.8 Smoking and vaping

Smoking is still a preventable killer in the UK. As well as the human cost, smoking costs the UK economy £14.7 billion per year, £2.5 billion of which falls to the NHS (Public Health England, 2018). Smoking kills people of working age as well as older people (Action on smoking and health, 2022).

Cost-effectiveness and cost-utility analyses were conducted in the Netherlands comparing a work-based smoking cessation training programme with and without incentives (van den Brand et al., 2020). The training programme was deemed cost-effective at a societal willingness to pay of €11,546 per additional quitter. The cost-effectiveness acceptability curve demonstrated an 80% probability of the intervention being cost-effective at €20,000 per abstinent smoker (van den Brand et al., 2020). Secondary cost-utility analysis revealed similar QALYs between the intervention and control groups, therefore it is not likely to be cost-effective (van den Brand et al., 2020). In a separate study conducted in Korea, a workplace smoking cessation programme yielded a return on investment of $15.39 for every $1 invested in the programme (Kim et al., 2022).

The NHS has advocated vaping (the use of e-cigarettes) as a strategy to help people quit tobacco smoking through the implementation of a ‘swap to stop’ scheme to encourage one million smokers to switch from tobacco cigarettes to vapes (Health and Social Care, 2023). This world-first national scheme provides smokers in England with a vaping starter kit and additional behavioural support to help meet the government goal of becoming smoke free (reducing the rates of smoking among the population to 5% or less) by 2030 (Health and Social Care, 2023). An economic modelling study conducted in New Zealand assessed the health and costs of electronic nicotine delivery systems compared to conventional smoking (Summers et al., 2022). The findings indicated that the liberalisation of electronic nicotine delivery systems could produce cost saving QALY gains across the lifespan of the New Zealand population (Summers et al., 2022). In the USA, a survey of 1,607 working age adults was conducted to explore issues relevant to employers with respect to the impact of vaping in the workplace (Graham et al., 2020). The survey findings revealed that workplace vaping was a trigger for smoking and vaping among current tobacco smokers. Observing workplace vaping was also found to be a trigger for smoking among 7% of former tobacco and vaping users (Graham et al., 2020). In addition, child vaping was also linked to presenteeism and absenteeism among approximately one third of parents due to feelings of anxiety and worry about their children’s use of vapes (Graham et al., 2020). A substantial proportion of the sample (84.8%) reported that a workplace that supports e-cigarette cessation was important to them (Graham et al., 2020). Research suggests that vaping is far less harmful than smoking tobacco, but the long-term health harms of vaping remain largely unknown (Hamberger & Halpern-Felsher, 2020; Hawk & Maresso, 2019). See Table 6 for studies relating to smoking.

**Table 6.** Economic studies relating to smoking cessation.

| Author | Date and Country | Aim | Intervention | Length of follow-up | Type of study | Population | Findings | Type of economic analysis and main finding | *Quality rating |
| --- | --- | --- | --- | --- | --- | --- | --- | --- | --- |
| Kim et al. | 2022<br>South Korea | To evaluate a workplace smoking cessation group training programme with incentives compared with a training programme without incentives. | A smoking cessation group training programme with incentives. | Unclear. | ROI. | N=82. | The workplace smoking cessation program led to 40.4% of participants quitting smoking and improvements in attitudes and perceptions related to smoking cessation. There were workplace-level reductions in productivity losses and medical expense losses. These economic effects were robust in the sensitivity analysis. In the future, it would be beneficial to conduct an economic evaluation to encourage the voluntary involvement of workplaces when implementing workplace health promotion programs. It would be also important to develop multi-level approach programs and expand its application. | ROI.<br>The mean ROI was 15.06 (minimum of 1.00 and maximum of 43.63) and considering both reductions in productivity loss and medical expenses, the mean ROI was 15.39 (minimum of -1.00 and maximum of 44.53). This means that the net benefit was about 15.06 and 15.39 times the cost, respectively. | High |
| Van den Brand et al. | 2019<br>The Netherlands | To evaluate a workplace smoking cessation group training programme with incentives compared with a training programme without incentives. | A smoking cessation training programme. The training consisted of seven sessions of 90 minutes each, distributed over a period of 2 months. | 14 months. | Cost-effectiveness analysis and cost-utility analysis. | N=604 | Of the participants in the intervention group, 41% had quit smoking compared with 26% in the control group. | Cost-effectiveness analysis.<br>There was no economic difference between the intervention and control group within the 14-month follow-up period. | High |

\*The Joanna Briggs Institute (JBI) tool used to rate the quality of the studies is designed for full economic evaluations and may have disadvantaged the quality rating of partial economic evaluations such as return on investment analyses.

### 17.9 Healthy eating and physical activity

A cost-effectiveness analysis of the ‘Fuel Your Life’ worksite weight management programme produced positive economic benefits (Corso et al., 2018). The economic evaluation compared three intervention conditions: telephone coach, small group coach, and self-study. In terms of weight loss, the small group intervention was more costly and was similar in effectiveness when compared to the self-study intervention. In comparison to the self-study group, the small group coach was found to be cost-effective at an acceptable range of $22,400 per QALY (Corso et al., 2018).

Two economic evaluation studies have reported favourable findings in relation to the cost-effectiveness of workplace sugar control (Basu et al., 2020) and system-level dietary modification ‘Food Choices at Work’ (Fitzgerald et al., 2018). Sugar is known as a key factor in obesity and people who have a higher body mass index (BMI) tend to eat and drink more sugary foods and drinks than those with a lower BMI. It was found that a ban on sugar sweetened beverage sales in the workplace could be cost saving from the employer perspective due to productivity expenditure averted. The mean saving per 10,000 employees over 10 years was $398,949 (Basu et al., 2020).

Scoping searches identified four studies of physical activity workplace initiatives with economic outcomes. Of these, two were economic evaluations exploring the cost-effectiveness of a telehealth worksite exercise intervention to reduce lost work time due to lower back pain among firefighters (Dagenais et al., 2021), and a loyalty scheme for physical activity behaviour change maintenance (Hunter et al., 2018). The third study was a costing analysis of an incentivised workplace physical activity intervention to increase daily steps among inactive employees (Mason et al., 2018). The fourth paper was a systematic review assessing the economic impact of workplace physical activity interventions in Europe (Braun et al., 2022).

Scoping searches identified four reviews assessing the cost-effectiveness of interventions to reduce occupational sitting time at the workplace (Akhavan Rad et al., 2022; Gao et al., 2019; Lutz et al., 2020; Nguyen et al., 2022). Searches also uncovered four full economic evaluations that provided supporting evidence on the cost-effectiveness of interventions to reduce occupational sitting time (Ben et al., 2022; Gao et al., 2018; Michaud et al., 2022; Munir et al., 2020). See Table 7 for studies relating to healthy eating and physical activity.

**Table 7.**
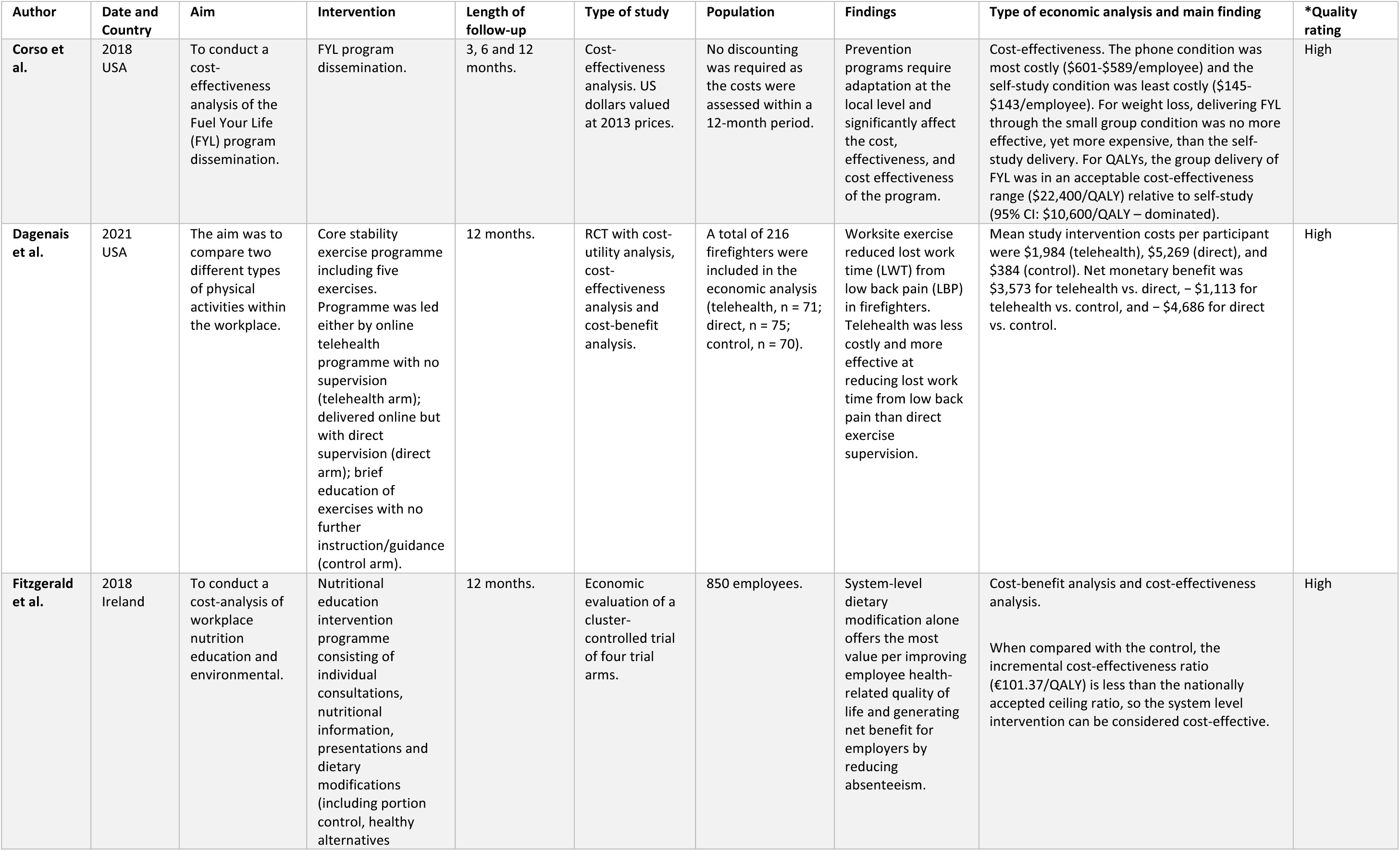
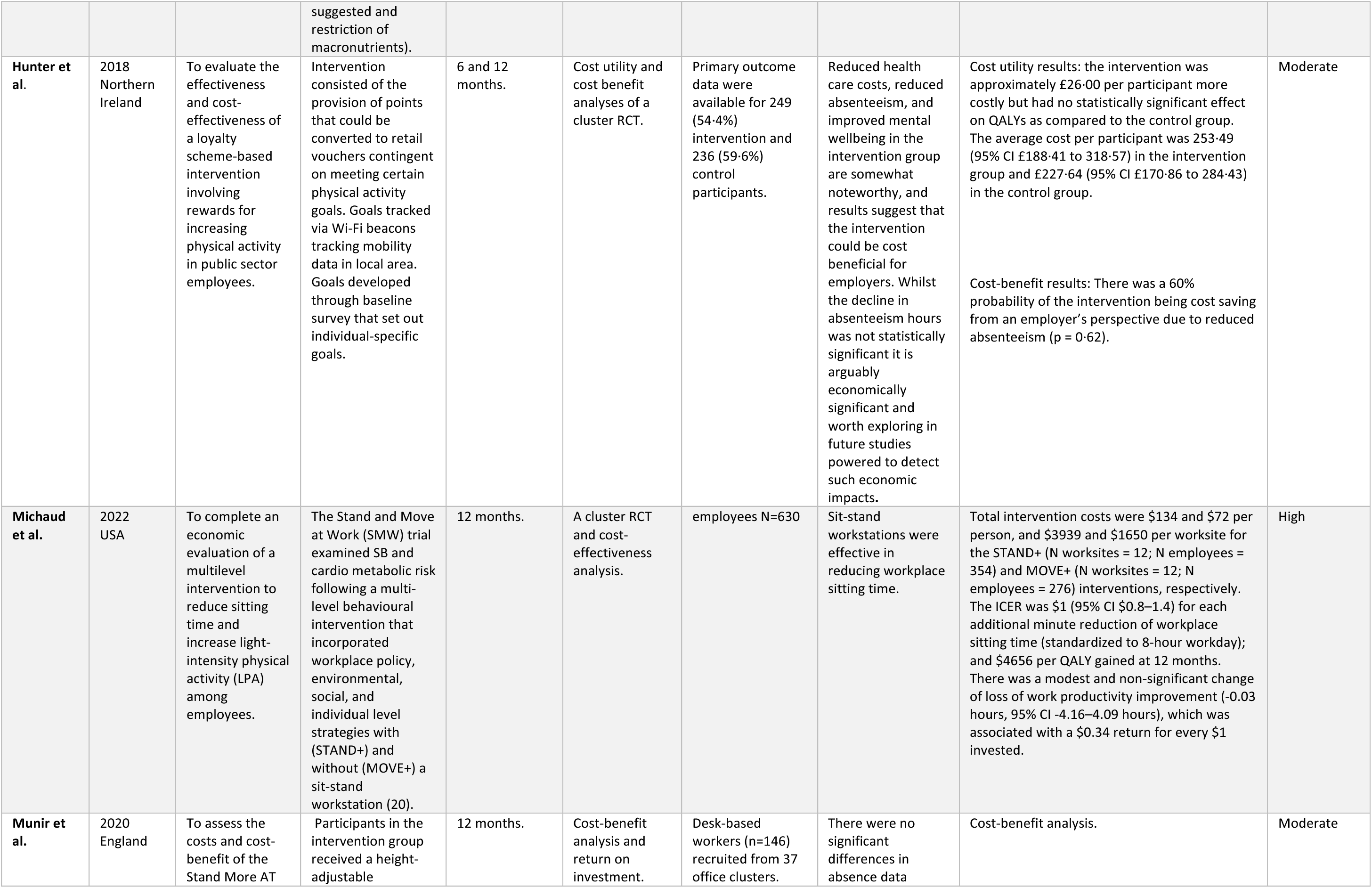
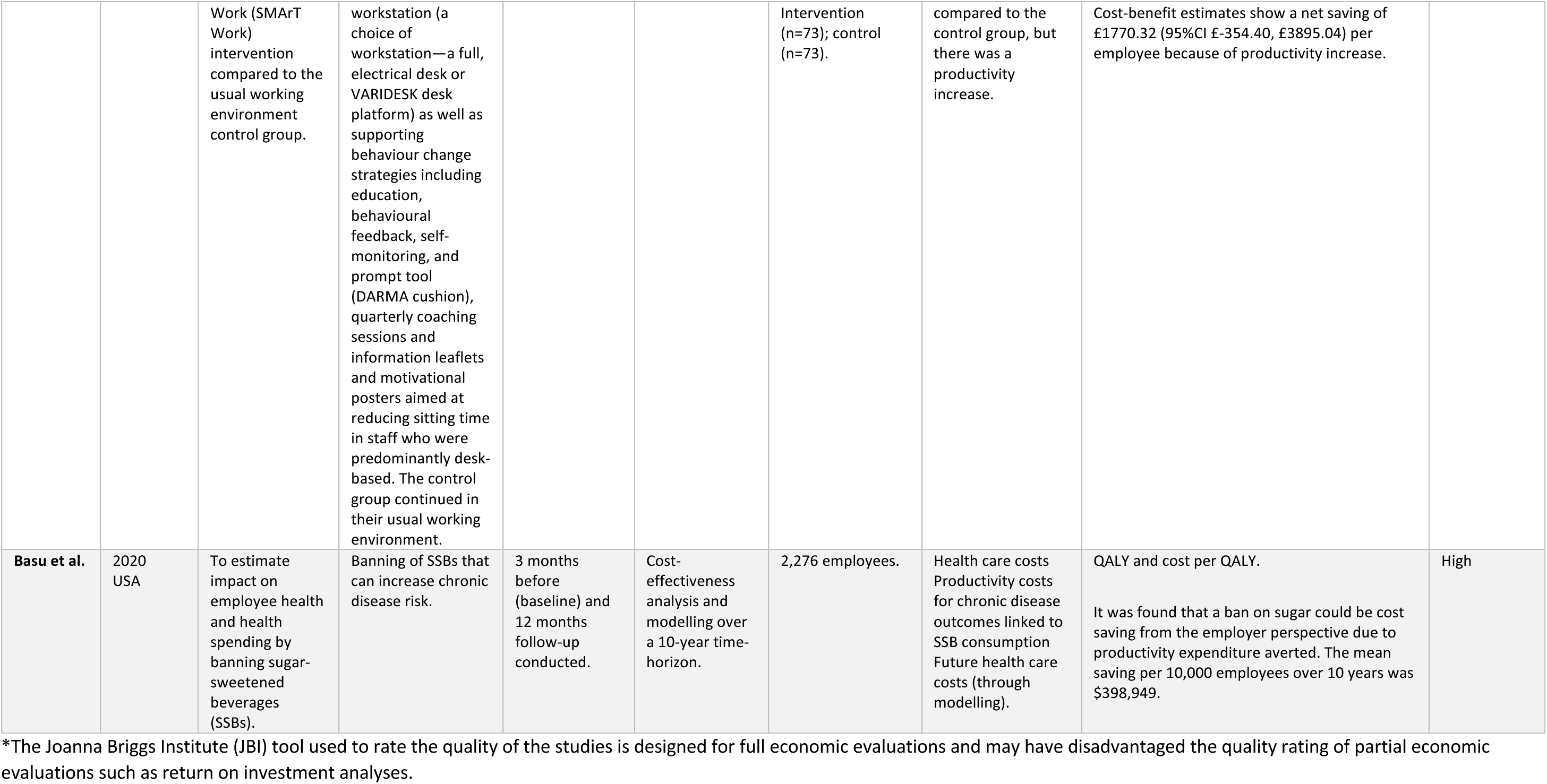
Economic studies relating to healthy eating and physical activity.

## 18 A spotlight on the health and social care workforce

### 18.1 Economic evidence concerning the health and social care workforce

The health and social care workforce are the largest in the Welsh economy, employing over 180,000 individuals. A substantial proportion of the health and social care workforce are women, and the workforce is aging. Nearly 40% of the health and social care workforce were aged over 50 in 2020, a gradual increase from just 29% in 2009. As of 2020, 41% of the Welsh health and social care workforce worked on a part-time basis. Interestingly, the proportion of women working part-time (48%) is significantly higher than men (15%) (Social Care Wales, 2020).

As demands on the NHS in Wales rise, workload pressures on health and social care workers increase. Burnout is intricately linked to recruitment and retention issues in the health care workforce (Galleta-Williams et al., 2020). A cross-sectional survey of 232 General Practitioners (GP) in the UK found that 93.8% of respondents were categorised as ‘likely suffering from a minor psychiatric disorder’ and 94% were experiencing mild (22%) or severe (74%) exhaustion (Hall et al., 2019). Research has indicated that presenteeism may be linked to job burnout caused by inadequate recovery from illness as workload pressures continue to rise (Homrich et al., 2020). Nurses and physicians are more prone to presenteeism compared to other professional groups, which in turn negatively impacts health outcomes, productivity, and costs (Lui et al., 2018). A systematic review has highlighted a lack of economic evaluations exploring presenteeism among hospital nurses and physicians (Lui et al., 2018). In terms of the available published economic evidence in this area, scoping searches have indicated that costing analyses of workplace well-being interventions have focused on the costs of depression-related presenteeism among resident physicians (Rosen et al., 2018), osteoporosis screening for healthcare workers (Rinaldi et al., 2021), and behaviour change strategies to reduce hand dermatitis among NHS nurses (Madan et al., 2020).

In Spain, an economic evaluation was conducted from a health system and societal perspective to explore the cost-effectiveness of a complex intervention to prevent and manage musculoskeletal pain among nurses (Soler-Font et al., 2019). At 12-months follow-up, the intervention was found to be cost-effective in reducing neck, shoulder, and upper back pain among nurses (Soler-Font et al., 2019).

Dealing with preventable health issues, unhealthy behaviours, and reducing the risk of injuries may decrease premature mortality and keep many working people at work for longer (Hussein et al., 2016). Staff retention is an important aspect of cost-effectiveness of the workforce. Improving workforce well-being is important in reducing presenteeism and sickness absence (Hussein et al., 2016). The act of listening alone can be beneficial to staff members. Listening has been identified as a key workplace skill, important for ensuring high-quality communication, building relationships, and motivating employees (Kriz et al., 2021).

#### 18.1.1 Health promotion

Public Health Wales launched the Healthy Working Wales programme to meet the workplace demands of all sectors across Wales. Such domains include healthy working environments, healthy lifestyle behaviours and person-centred leadership, which supports employees with health conditions (Public Health Wales, 2023b). Health promotion interventions are usually targeted and responsive to changing workforce needs.

## 19 Discussion

Our *Wellness in work* report is set within the context of the wider UK economic landscape and trends in premature morbidity and mortality. The UK is forecast to experience five years of lost economic growth, the longest since the global financial crisis of 2008 (National Institute of Economic and Social Research, 2022).

This report has considered 1) the wider economic impact of well-being and well-becoming in the workforce, 2) economic evidence of interventions that promote well-being and well-becoming to enable retention and return to the workforce.

With rising inflation and a persistent cost of living crisis affecting the UK, many employees are experiencing poor mental health due to worrying about their finances. 60% of people in Wales agreed that rising costs of living negatively affected their quality of life (25% strongly agreed). 87% reported ‘worrying’ about the cost of living, with 38% reporting ‘worrying a lot’ (Public Health Wales, 2023b). People with poor mental health are more likely to experience subsequent reductions in income. The negative impacts on mental health and well-being induced by the cost of living crisis could lead to future financial and health problems, creating an entrenched downward spiral that can persist even if economic conditions do improve (Thomson et al., 2022).

Staff well-being is an important factor in workplace productivity. Increased employee well-being is associated with lower levels of absenteeism and sick leave, greater resilience, better staff retention and increased staff commitment and productivity (MindWell, 2023). Universal and inclusive primary prevention initiatives such as health promotion activities and organisation wide culture change can have a high positive return on investment and empower staff to thrive at work. Flexible working policies available to all staff can help people with caring responsibilities to re-join or maintain employment. In recent years, there has been a shift towards hybrid working in many workplaces as a result of the COVID-19 pandemic. Many people in Wales continue to follow a hybrid working pattern (Chwarae Teg, 2022).

By 2040, 1 in 5 people (19%) aged 20 years and older are projected to be living with a major illness, moving from almost 1 in 6 in 2019 (Watt et al., 2023). This Wellness in work report did not identify evidence on the cost-effectiveness of ‘fit note’ schemes to help people return to work, but this could be a focus of future research (Public Health Wales, 2023a). Targeted interventions within workplaces are required to support the well-being of staff who are at risk of struggling or who are struggling with poor physical and mental health. The reduction of sickness-related absence could facilitate returning to and remaining in the workforce. Promoting employee mental well-being will produce important net economic benefit and produce financial gains, of the order of £100 million per annum for the UK (National Institute for Health and Care Excellence, 2022b).

Due to an ageing population in Wales, people are working later in life and combining work and caring responsibilities. Supporting older people to stay in the workforce for longer can generate productivity gains through retaining cumulative experience and skills in the workplace (ILC, 2022).

Tackling the lower levels of employment of women and ethnic minorities in Wales is critical for individual well-being and the Welsh economy. Since 2019, 16,000 women have left employment in Wales. During the same period, 13,000 men have gained employment (Welsh Government, 2023g). The employment rate for working-age ethnic minorities in Wales was 68% in 2022. Comparatively, the employment rate of working-age white individuals was 74% in 2022.

People with disabilities are less represented in the workforce than non-disabled individuals both at a UK and Welsh level. In 2022, 49% of people in Wales with a disability were in paid employment, significantly less than the 74% of non-disabled individuals (Welsh Government, 2023g). Interventions targeting people living with short and long-term disabilities, as well as neurodiverse people to join and stay in the workforce, could increase inclusivity, and bring economic benefits to the Welsh economy.

Many of the economic evaluations included in this report found that there were monetary benefits to interventions such as standing desks, healthy eating programmes, provision of healthy eating information, over the phone counselling, mental health training and gentle physical activity interventions. Benefits to the employer arose from reduced absenteeism and productivity increases. Wider societal benefits manifested in reduced burden on the healthcare system and increased employment levels. However, other studies showed no economic benefits of interventions. For example, a smoking cessation intervention did not result in changes in productivity costs to the employer but could result in health and social benefits for the employee.

Consideration must be given to the time horizons in the studies reported. The NICE reference case recommends using a time horizon long enough to reflect all important differences in costs or outcomes between the interventions being compared (National Institute for Health and Care Excellence, 2022c). Many of the time horizons identified in this review were very short (generally no more than 12 months), and this was especially true for drug and alcohol-related workplace interventions. This is a limitation of the evidence identified.

In drawing the conclusions together from this report, prevention of avoidable ill-health, disability, and premature death is the responsibility of all of us, including employers and employees, through co-produced better health.

This report aligns with the Welsh Government’s Wellbeing of Future Generations Act and their Prosperity for All national strategy as well as the UK Wellness Workforce plan. Through the Wellbeing of Future Generations Act Wales is seeking a more equal, prosperous, resilient, healthier, and globally responsive Wales.

> Taking care of the workforce is important for productivity as healthy and happy employees are more able to thrive and remain in the workforce for longer. Promotion of flexible working should be considered for employees who express an interest or preference for such work.

## 20 Glossary

**Absenteeism** – The time an employee spends away from the workplace. Absences can be scheduled (e.g., annual leave) or unscheduled (e.g., due to injury or illness).

**Alcohol misuse** – Alcohol consumption that puts individuals at increased risk for adverse health and social consequences.

**Brexit -** Brexit was the withdrawal of the United Kingdom from the European Union on 31 January 2020. The rules governing the new relationship between the EU and UK took effect in January 2021.

**Body Mass Index (BMI)** – A number calculated from a person’s weight and height; a high BMI score can lead to health problems.

**Burnout** – Defined by the International Statistical Classification of Diseases and Related Health Problems as a “state of vital exhaustion.” P.227

**Cognitive Behavioural Therapy (CBT)** – A type of psychotherapy in which negative patterns of thought about self and the world are challenged to alter unwanted behavioural patterns or treat disorders such as depression.

**Communities for Work Wales -** Different counties of Wales have their own communities for work programmes, backed by European Funding. According to the Welsh Government, £135 million has been invested into the Communities for Work and Parents, Childcare and Employment schemes since 2015 through EU structural funds, with more than 17,500 helped into employment.

**Communities for Work Plus** - Provides specialist employment advisory support and intensive mentoring to people who are under-represented in the labour market including young, old, and disabled people; Black, Asian, and Minority Ethnic people; and those with care responsibilities.

**COVID-19** – Coronavirus disease (COVID-19) is an infectious disease caused by the SARS-CoV-2 virus.

**Economically active** – People who are either in employment or unemployed.

**Economic activity rate** - People, who are economically active, expressed as a percentage of all people.

**Economic inactivity** - People who are neither in employment nor unemployed. This group includes, for example, all those who were looking after a home or retired.

**Employment rate** - The number of people in employment expressed as a percentage of all people aged 16-64.

**Employees and self-employed** - The division between employees and self-employed is based on survey respondents’ own assessment of their employment status.

**Endometriosis** – Endometriosis is a disease in which tissue like the lining of the uterus grows outside the uterus. It can cause severe pain in the pelvis and make it harder to get pregnant.

**Equity** - The term “equity” refers to fairness and justice and is distinguished from equality: Whereas equality means providing the same to all, equity means recognizing that we do not all start from the same place and must acknowledge and adjust imbalances.

**Furlough –** Furlough is a Job Retention Scheme recently used during the COVID-19 pandemic.

**Gender pay gap** – The average difference between pay levels for men and women who are working. Gross Domestic Product (GDP) – The Gross Domestic Product measures the value of economic activity within a country. GDP is the sum of the market values, or prices, of all final goods and services produced.

**Gig worker** - A gig worker is a person who participates in short-term contracts or freelance work. This can include anyone from Uber drivers to freelance writers. This kind of employment is good for people who want to work flexibly.

**Green economy -** A green economy is an economy that aims at reducing environmental risks and ecological scarcities, and that aims for sustainable development without degrading the environment. It is closely related with ecological economics but has a more politically applied focus.

**Gross Value Added (GVA)** – GVA measures the contribution to the economy of each individual producer, industry, or sector in the UK. It is used in the estimation of gross domestic product (GDP).

**Households** - A household is defined as a single person, or a group of people living at the same address who have the address as their only or main residence.

**Horizontal segregation** – The fact that there are more men than women doing one type of job and more women than men doing another type of job.

**Intervention** – A generic term used in public health to describe a policy or programme designed to have an impact on a health problem.

**In employment** - People who did some paid work in the reference week (whether as an employee or self-employed); those who had a job that they were temporarily away from (e.g., on holiday); those on government-supported training and employment programmes; and those doing unpaid family work.

**In-work poverty** – Low pay is a trigger for in-work poverty, which it is more likely to occur when only one adult in the household is working in paid employment.

**Jobs Growth Wales Plus -** Jobs Growth Wales+ is a training and development programme for 16–19-year-olds that provides skills, qualifications and experience young people need to get a job or further training.

**Job Retention Scheme** – Job retention schemes tend to be temporary schemes designed to protect the UK economy by helping employers whose operations may be affected by outside influences. An example of a job retention scheme is the furlough scheme utilised by the UK Government during the COVID-19 pandemic.

**Just About Managing (JAM) families** - Families that are not rich, but also not the poorest in society, and who despite mostly being in work, find day-to-day life a struggle.

**Labour supply** - Labour supply consists of people who are employed, as well as those people defined as unemployed or economically inactive, who can be potential labour supply.

**Long-COVID** - The World Health Organisation (WHO) defines Long COVID as “the continuation or development of new symptoms 3 months after the initial SARS-CoV-2 infection, with these symptoms lasting for at least 2 months with no other explanation.”

**Lost Productivity Time (LPT)** – Absence and reduced performance of the workforce resulting in reduced profits or benefits for the employers.

**Menopause** – The menopause is when a woman stops having periods. This is a natural part of ageing that usually happens between 45 and 55 years old.

**Not in Education, Employment, or Training** (NEET) – NEET is a term used in the UK to describe a young person who is no longer in the education system and who is not working or being trained for work.

**Not wanting a job** - People who are neither in employment nor unemployed and who do not want a job.

**No Guaranteed Hours Contracts (NGHCs)** – A type of work contract where there is no guarantee of any hours of work per week (also known as zero hours contracts in the UK).

**Occupation** - Occupations are classified according to the Standard Occupation Classification 2010.

**Opportunity cost** – The value of benefits foregone by not using resources in their next best alternative use.

**Presenteeism** – The measurable extent to which health symptoms, conditions and diseases adversely affect the work productivity of individuals who choose to remain at work.

**Primary prevention initiatives** – aim to prevent disease or injury before it ever occurs.

**Pro-rate** – To divide, distribute, or assess proportionately.

**Productivity** – A measure of worker output impacted by the worker’s health status.

**Quality Adjusted Life Year (QALY)** – This is defined as a year of life adjusted for its quality of life. Patients may gain added years of life from a treatment or intervention. This time is adjusted by the quality of life during that period.

**ReAct+ -** ReAct+ offers tailored solutions which may include financial support, skills training, and Personal Development Support to help remove barriers to employment, such as support with mental health, confidence building, language skills and more.

**Return on Investment (ROI)** – This is the net economic return for each pound invested in a public health intervention. It is expressed as either a percentage, or it can be stated that each £1 invested will generate e.g., £7.10 in economic returns. The £7.10 does not include the original £1 invested.

**Return to work (RTW)** – Returning to employment after a period of absence from work.

**Secondary prevention** – trying to detect a disease early and prevent it from getting worse.

**Sickness absence/sick leave** - Employees can take time off work if they are ill. They need to give their employer proof if they are ill for more than 7 days.

**Small and medium-sized enterprises (SME)** – enterprises with fewer than 250 employees.

**Smoking cessation** – Stopping or quitting using tobacco. Methods include counselling or medications to stop tobacco use. Social capital – The social glue that helps people, organisations, and communities to work together towards shared goals.

**Social Return on Investment (SROI)** – Social Return on Investment (SROI) is a framework for measuring and accounting for this much broader concept of value; it seeks to reduce inequality and environmental degradation and improve well-being by incorporating social, environmental and economic costs and benefits.

**Staff retention** - Staff retention is the ability of an organization to retain its employees and ensure sustainability.

**Sustainable work** - Sustainable work means achieving living and working conditions that support people in engaging and remaining in work throughout an extended working life. Work must be transformed to eliminate the factors that discourage or hinder workers from staying in or entering the workforce.

**Tertiary Prevention** – trying to improve your quality of life and reduce the symptoms of a disease you already have. Unexplained portion of the pay gap – Some of the pay gap between men and women can be attributed to known factors such as age, education and the type of jobs men and women tend to do, but there is a substantial portion of the pay gap which remains unexplained.

**Thriving employees –** Thriving employees are motivated by their work and experience great personal growth on the job. They have a positive psychological state and experience both a sense of vitality and learning.

**Unemployed -** Unemployment refers to people without a job who were available to start work in the two weeks following their interview and who had either looked for work in the four weeks prior to interview or were waiting to start a job they had already obtained.

**Unemployment rate** - Unemployed as a percentage of the economically active population.

**Upper limb disorders** – Upper limb disorders (ULDs) affect the arms, from fingers to shoulder, and neck. They are often called repetitive strain injuries (RSI), cumulative trauma disorder or occupational overuse syndrome.

**Vertical segregation** – The situation where people do not get jobs above a particular rank in organizations because of their race, age, or sex: Career progression of women and men in the higher education sector confirms a pattern of vertical segregation. Women often reach a ‘glass ceiling’ in careers due to vertical segregation.

**Waiting lists** – Elective and emergency treatment lists in the NHS.

**Wanting a job** - People not in employment who want a job but are not classed as unemployed because they have either not sought work in the last four weeks or are not available to start work.

**Working from home (WFH)/Remote work** – The term remote work became popular during the 2020 to 2022 pandemic that forced most of the office and knowledge workers to work from home.

**Workforce -** The workforce is the people engaged in or available for work, either in a country or area or in a particular firm or industry.

**Workless households** - Households where no-one aged 16 or over is in employment. These members may be unemployed or economically inactive. Economically inactive members may be unavailable to work because of family commitments, retirement, or study, or unable to work through sickness or disability.

**Workplace health programmes** – A set of strategies which include programmes, policies, benefits, environmental supports, and links to the surrounding community designed to meet the health and safety needs of all employees.

**Work-related musculoskeletal disorders** – Injuries or disorders of the muscles, nerves, tendons, joints, cartilage, and spinal discs that work environments can make worse.

## Data Availability

All data produced in the present study are available upon reasonable request to the authors

## 21 Acknowledgements

We wish to acknowledge the support of Dr Catherine Lawrence for critical feedback and proofreading, and Dr Sofie Roberts for proofreading. We would like to thank our stakeholders, Emma Edworthy, Melda Lois Griffiths, Pauline Mould, Samantha Huckle, Olivia Gallen and Libby Humphris for their time, expertise and input.

## 22 Author statement

Conceptualisation: RTE. Writing: RTE, LHS, BFA, AM, KP, JD, HL-W and DF. Review and editing: RTE, DF, LHS, BFA, JD, KP, AM, SB, AC, BC, JMC, RL, and MM.

## 24 List of appendices

### 25 Appendix 1. PICO table

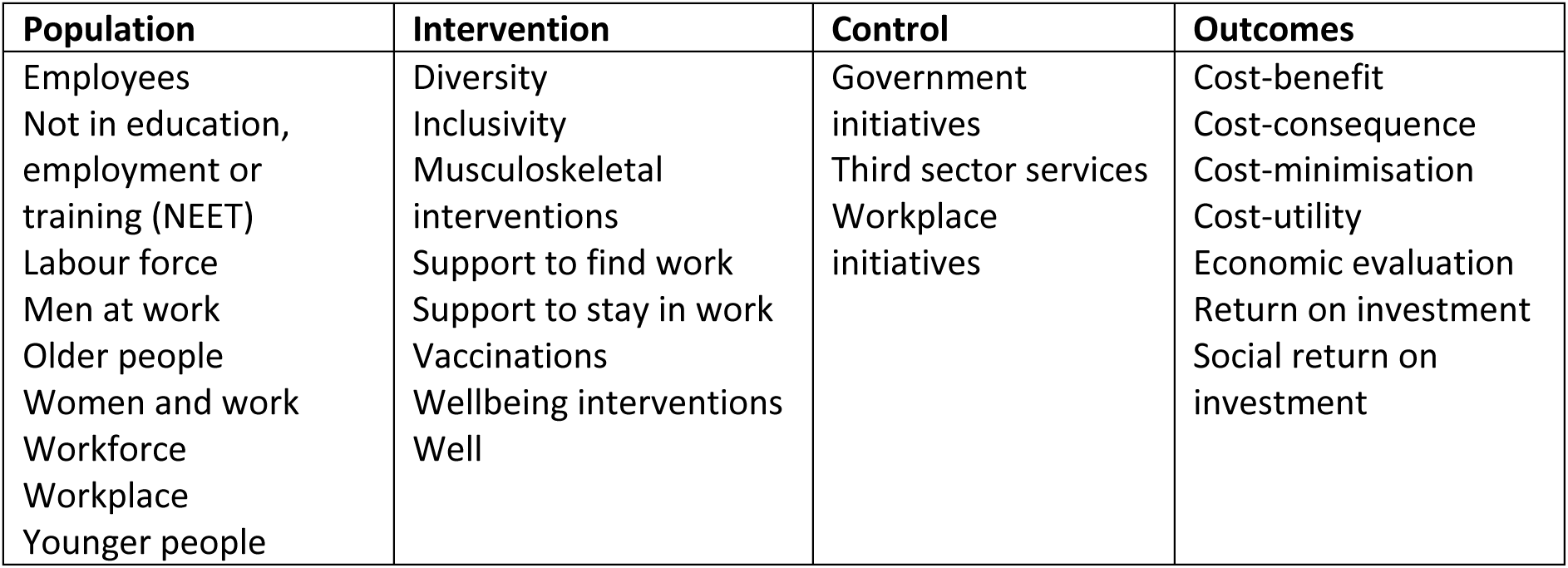

| Population | Intervention | Control | Outcomes |
| --- | --- | --- | --- |
| Employees<br>Not in education, employment or training (NEET)<br>Labour force<br>Men at work<br>Older people<br>Women and work<br>Workforce<br>Workplace<br>Younger people | Diversity<br>Inclusivity<br>Musculoskeletal interventions<br>Support to find work<br>Support to stay in work<br>Vaccinations<br>Wellbeing interventions<br>Well | Government initiatives<br>Third sector services<br>Workplace initiatives | Cost-benefit<br>Cost-consequence<br>Cost-minimisation<br>Cost-utility<br>Economic evaluation<br>Return on investment<br>Social return on investment |

### 26 Appendix 2. Search strategy performed in Medline via Ovid

1. Workplace/
2. well-being at work.mp.
3. (work adj3 well-being).mp.
4. (well-being adj3 employment).mp.
5. (healthy adj3 workplace).
6. (health* adj3 workplace).mp.
7. (health* adj3 employment).mp.
8. (healthy adj3 employment).mp.
9. (well-being adj3 employee).mp.
10. 1 or 2 or 3 or 4 or 5 or 6 or 7 or 8 or 9
11. Cost-Benefit Analysis/ or cost-effective.mp.
12. cost-benefit.mp.
13. cost-utility.mp.
14. cost-consequence.mp.
15. cost-minimisation.mp.
16. cost-minimization.mp.
17. social return on investment.mp.
18. SROI.mp.
19. return on investment.mp.
20. economic evaluation.mp.
21. 11 or 12 or 13 or 14 or 15 or 16 or 17 or 18 or 19 or 20
22. 10 and 21

### 27 Appendix 3. PRISMA flow diagram

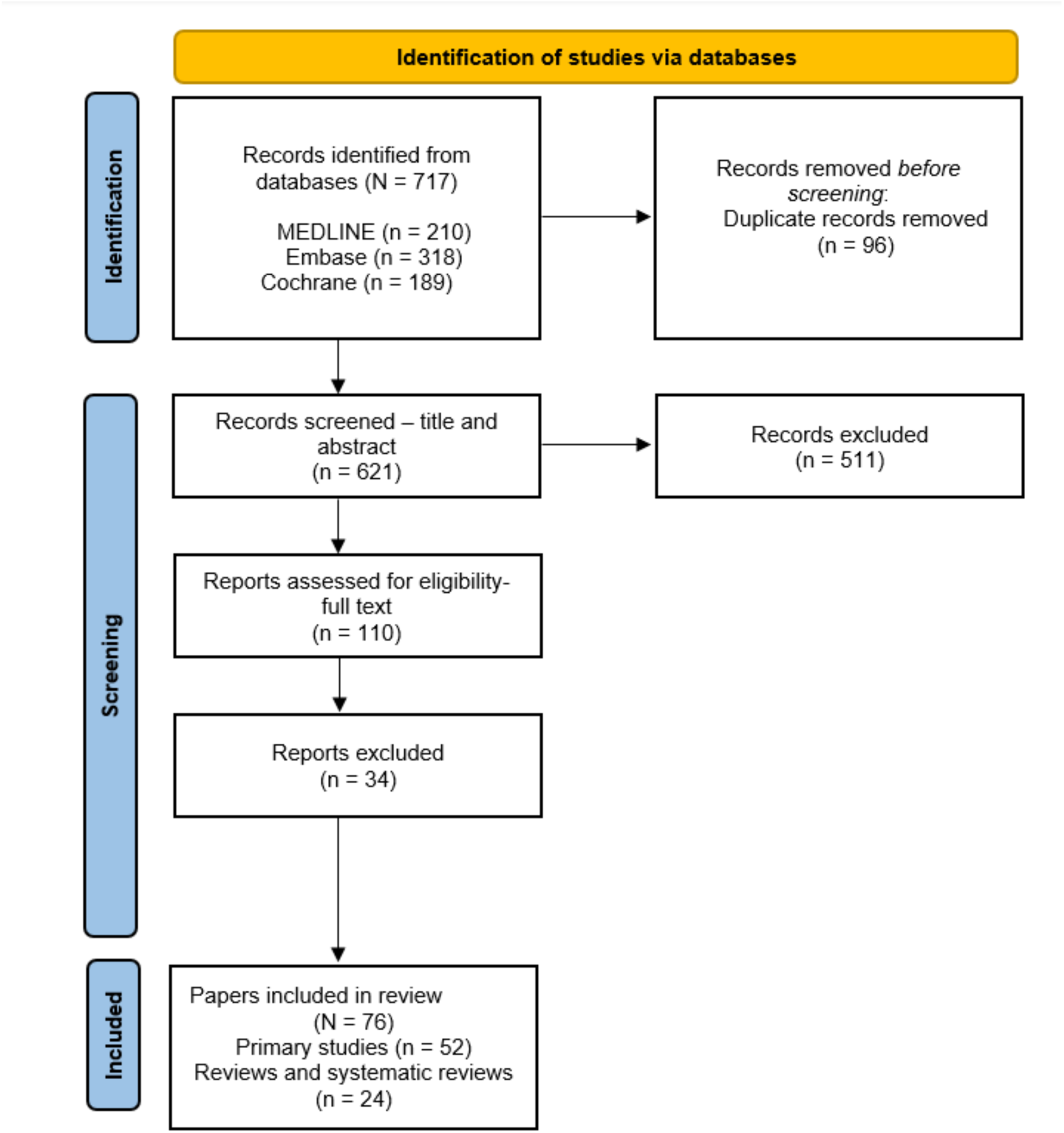
PRISMA flow diagram (Page et al., 2021).

### 26 Appendix 4. Data extraction tables

This is available on request to the Health and Care Research Wales Evidence Centre.

### 27 Appendix 5. Summary statistics regarding the Welsh workforce

**N.B: The figures used below were obtained from the Annual Population Survey 2022 and Labour Market Statistics February 2023 release. More up to date sources are used in the text. These summary statistics were sent to stakeholders on Monday 22^nd^ May 2023.**

The COVID-19 pandemic, exit of the United Kingdom (UK) from the European Union (also known as Brexit) and a significant cost of living crisis have changed the health and well-being of the labour force in Wales (Dalingwater, 2019; Edwards et al., 2019; The Kings Fund, 2022). Current trends of the Welsh labour force are offered below. The figures represent those of working age (16-64 years) unless otherwise stated and includes both part-and full-time employment patterns.

**The number of people employed in Wales has fallen since the start of the COVID-19 pandemic**

- There are 1.4 million individuals employed in Wales: a rate of 72%. A decrease of 6,000 or 2.3 Percentage Points (PP) from the 74% employment rate in 2019 (Welsh Government, 2023f, 2023h).
- There are 775,000 men in employment in Wales (78% of working age men). An increase of 5,000 since 2019 (Welsh Government, 2023f, 2023h).
- There are 666,000 women in employment in Wales (67% of working age women). A decrease in participation of 4.2PP from 2022. Since 2019, 26,000 women have left employment in Wales (Welsh Government, 2023f, 2023h).
- The employment rate for women in Wales is lower than the UK average and the gap between these rates has widened significantly from 0.8PP to 5.2PP in the year since 2022 (Welsh Government, 2023f, 2023h).
- Women aged 50-74 living in the ‘healthiest’ areas of England and Wales were 6% more likely to be in paid work than those living in the ‘unhealthiest’ areas (ILC, 2022).

**Overall unemployment has fallen in Wales since the start of the COVID-19 pandemic**

- 52,000 individuals are unemployed in Wales: a rate of 3.5%. This has fallen from 62,000 in 2019 (Welsh Government, 2023f, 2023h).
- There are 36,000 unemployed men, an increase of 10,000 (40%) from 2022. This has remained almost static from the 36,000 level in 2019 (Welsh Government, 2023f). This increase may relate to a greater dependency on self-employment, a sector that has exhibited both volatility and decline in number since 2020 (Office for National Statistics, 2023a).
- There are 17,000 unemployed women, a decrease of 3,000 (15%) from 2022. This decrease is a consistent trend observed from 2019, when 26,000 women were unemployed in Wales (Welsh Government, 2023f).

**Economic inactivity has increased significantly since the start of the COVID-19 pandemic**

- 476,000 individuals are economically inactive in Wales: a rate of 25%. This has increased by 28,000 or 2.3PP since 2022, the largest increase across UK nations. The Welsh economic inactivity rate has consistently been above the UK rate since 2019 (Welsh Government, 2023h).
- Of those economically inactive in Wales, 178,000 were men and 298,000 were women. Economic inactivity for men has fallen by 16,000 from 2022, indicating re-entry to the labour force (Welsh Government, 2023a).
- Conversely, the number of economically inactive women has increased by 44,000 since 2022 (31% of working age women). The number of women becoming economically inactive in the UK decreased in this same period. This may be partly explained by the dependence on health and social care jobs in the Welsh labour force, in which, women tend to be more represented (Welsh Government, 2023a).
- Looking after family or home (22%), being temporarily sick or injured (21%), retiring from paid work (18%) and being a student (12%) were the most common reasons for being economically inactive prior to reporting long-term sickness in the UK (Hickman et al., 2023).
- Increases in economic inactivity for women could be caused by but not limited to symptoms of menopause, caring responsibilities, unequal access to support and training, and historical gender-based inequalities in labour markets (Office for National Statistics, 2021d).
- At the UK level, increasing sickness, changes in migration structures, retirement patterns and an ageing population contribute to rising economic inactivity (House of Lords Economic Affairs Committee, 2023).

**The public sector continues to be a key employer within Wales**

- The public sector in Wales employed 321,000 people in 2022 (10% of total Welsh population). This figure increased by 8,000 (3%) from 2021 (Office for National Statistics, 2023e).
- Human health and social work activities accounted for the greatest number of workforce jobs in Wales in 2022 at 223,000 (15%) of the total workforce (Office for National Statistics, 2023e). In 2019, the figure stood at 214,000 highlighting a longitudinal dependence on the sector in Wales.

**Well-being is essential for ensuring a healthy and productive labour force. The workforce has been affected by long waiting lists for elective care exacerbated by the COVID-19 pandemic**

- Keeping people healthy and in work prevents loss of productivity and benefits the Welsh economy (Black, 2009).
- There is an 11-year gap in disability-free life expectancy (DFLE) between the healthiest and unhealthiest areas of England and Wales. Older workers from these areas are 60% more likely to be out of work than those from healthiest areas (Office for National Statistics, 2020). For those in the most deprived areas in Wales, women have an 18-year shorter healthy life expectancy than in the least deprived areas. For men, the figure is 17 years (The Health Foundation, 2022).
- Despite having one working adult in the household, 31% of children in Wales lived in poverty in 2022. An increase from 29% in 2019 (Welsh Government, 2019b, 2023c).
- The median gross weekly earnings for full time adults working in Wales was £598 in April 2022 (93% of the UK average of £640) (Welsh Government, 2022b). Furthermore, high inflation over the period starting winter 2021 has reduced real wages (Office for National Statistics, 2023a).
- Elective surgery waiting lists have reached 754,000 (just under 1 in 4 of the total Welsh population). Age breakdowns are not available in the data, but it is reasonable to argue a considerable amount of working age people are awaiting elective surgery that is limiting their ability to work or work productively (Welsh Government, 2023j).

**Place has a strong relationship to employment and entrenched deprivation persists in Wales**

- High employment deprivation persisted in numerous areas between the 2014 and 2019 Welsh Index of Multiple Deprivation (WIMD). High levels of employment deprivation exist in the South Wales Valleys, large Welsh cities, and coastal towns in North Wales (Welsh Government, 2019d).
- The percentage of people in income deprivation in Lower Layer Super Output Areas (LSOAs) in deep-rooted deprivation (43%) was almost 3 times that of areas that have never been ranked in the top 50 most deprived (15%). It was also approximately 3 times the Wales average (16%) (Welsh Government, 2022a).
- Some communities are highly reliant on anchor institutions for their employment. Mass unemployment events (localised economic shocks) can have adverse impacts on the health, financial and social circumstances of workers, families, and communities (Davies et al., 2019). The recent closure of the 2 Sisters Food Group factory in Llangefni, Anglesey resulted in the loss of 700 jobs in the local community (BBC Wales News, 2023).

**More individuals from ethnic minority communities are now in employment in Wales**

- The employment rate for working age ethnic minorities in Wales was 68% in 2022. A 3PP increase from 2021. Comparatively, the employment rate of working age white individuals was 74% in 2022, a 0.1PP increase on the previous year (Welsh Government, 2023h).
- The unemployment rate of working age ethnic minorities was 7% in 2022. A 3.9PP decrease from 2022. The unemployment rate for working age white individuals was 3% in 2022. A1.1PP decrease on the previous year (Powell & Francis-Devine, 2023).

**Less people living with disabilities are in employment in Wales than in other areas of the UK**

- The difference between disabled and non-disabled employment in Wales stands at 32.3PP. The UK figure is 29.8PP (Office for National Statistics, 2022a).
- Here in Wales, as of 2021, more working age disabled people had no qualifications than non-disabled people (13% vs 5%) (Office for National Statistics, 2022a).
- The employment patterns of disabled workers in Wales display a greater dependence on part-time work (Office for National Statistics, 2022b).
- Severe or specific learning difficulties and autism were the disabilities with the lowest employment rates of 26% and 29% respectively across the UK (Office for National Statistics, 2021b).

38,620 people are in receipt of Access to Work payments in the UK - a 2% increase from 2021. The most common element of receipt was Support Workers with 17,610 (making up 46% of all payments) (Department for Work and Pensions, 2022a).

